# Disrupting the Clock: Meta-Analysis of Irregular Night Shifts and Migraine, Proposing Shift Work Migraine Disorder with Chronobiology Strategies

**DOI:** 10.1101/2025.07.31.25332540

**Authors:** Yohannes W. Woldeamanuel, Ariana Rahman, Esam T. Hyimanot, Richa Chirravuri, Mahya Fani, Elika D. Javaheri, Madeline Welch, Joyce Zhuang, Chung Jung Mun

## Abstract

**Background:** Migraine is linked to circadian rhythm disruptions, with morning attack peaks, circadian variations in trigeminal pain sensitivity, anterior hypothalamus involvement, and core circadian clock gene activity. Irregular night shift work, affecting up to 50% of the population, including new parents and students, causes significant circadian disruption. We hypothesize that irregular night shifts increase migraine prevalence compared to fixed schedules.

**Methods:** A systematic review and meta-analysis of observational studies up to March 27, 2025, assessed migraine prevalence in irregular versus fixed night shift workers, searching Web of Science and PubMed with terms like “shift work” and “migraine” (PRISMA/MOOSE-compliant, PROSPERO: CRD420250654865). Study quality was evaluated using the Newcastle-Ottawa Scale (NOS). A random-effects meta-analysis calculated weighted odds ratios (ORs) for migraine prevalence.

**Results:** From 203 records, 13 high-quality cross-sectional studies (N=38,798,271, 77% female, NOS 9–10) showed irregular night shifts significantly increased migraine odds (OR=1.61, 95% CI: 1.27–2.04, p<0.0001, I²=73%), with females at higher odds (OR=2.02–4.21). Meta-regression linked higher female representation to increased migraine odds (β=0.70, p=0.0003, R²=50%). Irregular night shifts showed no association with tension-type headache (OR=0.79, 95% CI: 0.43–1.45).

**Conclusion:** Irregular night shifts disrupt circadian rhythms, elevating migraine odds but not tension-type headache, suggesting fixed schedules may reduce the burden. Chronobiology-informed management, including slow-rotating schedules (≥5 days with rest days), delay- directed rotations, timed light exposure, and ambient temperature regulation, needs testing to prevent ‘Shift Work Migraine Disorder,’ a proposed distinct migraine subgroup.

## Introduction

Migraine is a complex and debilitating neurological disorder characterized by recurrent episodes of severe headaches, often accompanied by sensitivity to light, sound, and nausea^1^. The pathophysiology of migraine is multifaceted, involving the interplay of various neural systems, including the trigeminal nerve, the brainstem, and the cerebral cortex^2^. Recent studies have highlighted the critical role of circadian rhythms in migraine pathophysiology, suggesting a significant link between disruptions in internal biological processes and migraine attacks^3–9^.

The observed circadian rhythmicity in migraine attacks exhibits distinct patterns, including matutinal/morning peaks in attack frequency, as well as circaseptan (weekly) and infradian (longer than 24 hours) patterns^4,7^. Furthermore, circadian variations in trigeminal pain sensitivity have been observed, suggesting a complex interplay between circadian rhythms and migraine pathophysiology^10^. The involvement of the anterior hypothalamus^11^, home to the suprachiasmatic nucleus (SCN), the "master clock" regulating circadian rhythms, and the expression of circadian-related genes such as CK1δ, PER2, and RORα, provide additional evidence supporting the link between circadian rhythms and migraine^4,12,13^.

The circadian system is a highly conserved and hardwired biological system that regulates various physiological processes, including pain sensitivity^14,15^, sleep-wake cycles^16^, neurotransmitter and hormone secretion^17,18^, and metabolism^17,19^. Disruptions to this intricate system, whether due to lifestyle factors, environmental influences, or genetic predispositions, can have far-reaching detrimental consequences, contributing to the development of various chronic diseases, such as cancer^20–24^, diabetes^19,25–27^, chronic pain^14,28^, cardiovascular disease^29^, and neurological disorders such as Alzheimer’s disease^30–32^.

Shift work involves organizing 24-hour operations into two or three distinct shifts, with start and end times varying based on shift length^33^. According to the US National Institute for Occupational Safety and Health (NIOSH), day shift typically runs from 5–8 a.m. to 2–6 p.m., evening shift from 2–6 p.m. to 10 p.m.–2 a.m., and night shift (colloquially known as “graveyard shift”) from 10 p.m.–2 a.m. to 5–8 a.m^33^. The International Labor Organization (ILO)^34^ and International Agency for Research on Cancer (IARC)^35^ define shift work as any work schedule outside conventional daytime hours, typically spanning 7:00 a.m. or 8:00 a.m. to 5:00 p.m. or 6:00 p.m., such as evening or night shifts. The ILO defines night work as any work performed during a period of at least seven consecutive hours, including midnight to 5 a.m., and a night worker as someone whose job involves a substantial amount of such hours exceeding a specified threshold^34^. The European Union (EU) Working Time Directive (WTD) adopts the same definition of "night time" as the ILO and defines a "night worker" as an individual who regularly works at least three hours of their daily shift during this period^36^. Globally, approximately 20% of the workforce engages in shift work, with regional variations, such as 12% in Europe (up to 58% in evening work)^37^ and 15% in Chile^38^, while in the U.S., shift work prevalence is highest in service industries like protective services (50.4%) and food preparation (49.4%)^38^. In other regions, night work affects 7.6% of workers in Brazil^39^, 16% in Australia^40^, 17.5% in China^41^, 20% in Senegal^42,43^, 21.8% in Japan^44^, and 28% in Canada^45^, with significant prevalence in sectors like healthcare, hospitality, manufacturing, and transportation. Approximately 27% of the U.S. workforce reports engaging in evening or night shift work, with 7.4% specifically reporting frequent night shift work, defined as working between 1:00 a.m. and 5:00 a.m. for 6 to 30 days within the preceding 30-day period^46^.

Migraine shows significant disparities in prevalence and severity across age groups^47,48^, sexes^48–50^, and races^51–56^. The specific impact of night shift work as a contributing factor to these disparities remains underexplored, warranting focused investigation.

- Age and Night Work Prevalence: According to the NIOSH, the prevalence of frequent night work demonstrates a clear age-related pattern, with the highest rates observed in the youngest demographic: 18-29 years (8.61%). This prevalence gradually decreases with increasing age: 30-44 years (7.78%), 45-64 years (6.93%), and is lowest in individuals aged 65 and older (3.68%)^46^. Similarly, as per the US Department of Labor (DOL) report, young workers (aged 15–24) are more likely to work non-daytime schedules, including evening, night, rotating, or irregular shifts, with 31.9% on such shifts compared to 16.4% of the total workforce^57^.
- Sex Differences: There was lower engagement in night work by females (5.57%) compared to males (9.11%)^46^. Studies show that females experience a migraine burden three times greater than males^48–50^. This increased susceptibility in females is further exacerbated by a higher prevalence of psychological comorbidities, such as anxiety and depression, and shorter free-running circadian cycles^58^. These factors may contribute to increased circadian disruption when subjected to night shift schedules, potentially amplifying the negative effects on migraine.
- Racial Disparities: Racial disparities in night work prevalence are also important. In the NIOSH report, Black individuals demonstrate the highest rates of night shift work (10.5%), followed by Whites (7.07%) and other racial groups (6.48%)^46^. The DOL report shows that Black workers are more likely to work non-daytime schedules, with 24.1% on such shifts compared to 15.2% of White workers and 16.4% of the total workforce^57^. Notably, research indicates that Black individuals tend to have shorter free-running circadian cycles compared to White individuals^58–60^. This physiological difference may further exacerbate the detrimental effects of circadian disruption associated with night shift work, potentially intensifying health disparities. Blacks also have limited healthcare access, lower treatment rates, and greater functional disability due to more severe migraine pain, intensifying migraine burden and disparities ^51–53,56,61,62^.
- Intersection of Demographics, Night Shift Work, and Migraine: The demographic profiles of younger females and Black workers align with populations already identified as having a higher prevalence and burden of migraine. Previous studies have consistently documented elevated migraine burden, increased severity, and pronounced disparities within these groups^48,50,53,55,61^. We hypothesize that frequent night work, with its inherent disruption of circadian rhythms, to be a significant trigger for both migraine exacerbation and de novo onset. Beyond ’traditional’ shift work, students facing academic pressure and new parents experiencing fragmented sleep due to newborn care are particularly vulnerable to night shifting and its associated circadian rhythm disruption^63,64^. Further in-depth investigation into the impact of night work on migraine prevalence and severity across all affected populations is warranted to develop targeted interventions and mitigation strategies that benefit diverse demographic groups.

Rotating or irregular night shift work, characterized by unpredictable changes in shift timing, is linked to greater circadian disruption, long sleep, depression, anxiety, and fatigue compared to fixed night shift work, likely due to increased recovery needs^65–71^. Rotating night shifts also significantly elevate psychological distress and impair sleep quality, posing substantial health risks^65,67^. Given the established link between circadian disruptions and migraine, we hypothesize that irregular or rotating night shift work, which is associated with increased circadian disruption, psychological distress, and reduced sleep quality compared to fixed night shift work, may correlate with a higher prevalence of migraine. Our objective is to investigate this relationship to inform evidence-based interventions aimed at reducing migraine burden in this vulnerable population.

## Methods

### Research Question

Our research question followed the PECO (Population, Exposure, Comparison, Outcome)^19^ format. Our study focuses on the general working population (P) and examines exposure to irregular night shift work or rotating night shifts with ≥ 5 night shift per month (E), defined by not remaining on the same night shift for at least two weeks. The comparison group (C) includes workers on regular non-rotating night and evening shifts or permanent night shift schedules, with <5 night shift days per month, while the outcome (O) of interest is the prevalence of migraine and headaches, measured using odds ratios from observational studies.

### Eligibility Criteria

The eligibility of studies was determined based on the components of the research question (PECO and study design). For inclusion, studies need to involve adult workers (aged 18 or older) in any occupation or industry, representing general working populations, not restricted to specific clinical or non-working groups (e.g., patients, retirees). Exposure was defined as frequent irregular or rotating night and/or evening shift work, characterized by frequent schedule changes (rotating or unpredictable night shifts)^34,36^. Comparison groups were required to include workers exposed to regular non-rotating night and evening shifts (fixed night schedules). Since evening and night shifts overlap (e.g., 9:00 p.m. to midnight), our study groups them together due to their shared characteristics and health impacts While there’s overlap in the hours (9:00 p.m. to midnight are included in both evening and night shifts^33,46,57^), both evening and night shifts are categorized together in these studies due to shared characteristics and health implications. Both shift types occur during peak melatonin production and sleep propensity, contributing to similar health risks^72^. Including both captures the broader impact of night shift work schedules on circadian disruption, despite overlapping hours.

Outcomes of interest were the prevalence of migraine, defined by clinical criteria (e.g., International Classification of Headache Disorders) or self-reported with validation, excluding other headache subtypes (e.g., tension-type or cluster headaches), or headaches when not subtyped (i.e., general or unspecified headaches, not categorized as migraine or other specific types). Studies were required to provide odds ratios (ORs) to quantify the association between shift work and migraine or headache prevalence, either directly reported ORs or data allowing for OR calculation (e.g., prevalence rates, contingency tables). Eligible study designs included observational studies, specifically cross-sectional studies, case-control studies, and cohort studies (prospective or retrospective), published in peer-reviewed journals or grey literature (e.g., theses, conference proceedings) with sufficient data, in any language (with translation available if needed), and with no date restrictions, given continuous knowledge updates. The 2015 NHIS report^46^ was included in our systematic review as it provided relevant data on night shift work and migraine, aligning with our study’s focus.

Exclusion criteria were applied to maintain focus and quality. Studies were excluded if they exclusively involved non-workers (e.g., students, unemployed individuals, retirees), were limited to pediatric populations (<18 years) or specific clinical cohorts (e.g., only migraine patients without a working context), lacked clear definitions of night shift work (e.g., no mention of timing or irregularity), did not differentiate shift work (e.g., combining day and night shifts without separate analysis), or focused solely on day shift work without night shift comparison. Studies without a comparison group (e.g., case series or descriptive studies with no control) or those comparing night shift work to irrelevant groups (e.g., unemployed individuals) rather than the specified comparators were also excluded. Additionally, studies were excluded if they did not report migraine or headaches as outcomes (e.g., focusing only on sleep disorders or fatigue), reported only headache subtypes other than migraine (e.g., tension-type or cluster headaches), unless headaches were non-subtyped, or provided only qualitative outcomes (e.g., no prevalence or OR data). Studies not providing odds ratios or sufficient data to derive them (e.g., only p-values or narrative results), using experimental or non-associational metrics (e.g., means without prevalence), or employing non-observational designs (e.g., randomized controlled trials, lab-based experiments) were excluded. Reviews, editorials, or opinion pieces without original data were also excluded, though their reference lists were screened for eligible studies. Studies with insufficient detail on shift work patterns (e.g., no frequency or quick return data) were excluded. Including studies where ORs could be calculated from prevalence or contingency tables (not just reported) broadened the pool of eligible studies without sacrificing rigor. All reviewers used these criteria to screen titles and abstracts, then full texts, resolving discrepancies through consensus. The criteria ensured that studies aligned with the question’s focus on the impact of irregular night shifts on migraine and headaches, while excluding irrelevant or low-quality data.

### Search Strategy and Study Selection

The process for identifying, screening, and selecting studies followed the Preferred Reporting Items for Systematic Reviews and Meta-Analyses (PRISMA) guidelines. The search strategy was tailored to the research question and the inclusion/exclusion criteria, specifying the databases, keywords, and time frame used to identify observational studies examining the association between irregular or rotating night shift work and the prevalence of migraine or non-subtyped headaches among workers, as measured by odds ratios or calculable odds ratios (e.g., from prevalence or contingency tables). The databases searched included PubMed and Web of Science. The search used a combination of controlled vocabulary (e.g., MeSH terms in PubMed) and free-text keywords to capture the population, exposure, comparison, outcome, and study design elements, grouped by concept and combined using Boolean operators (AND, OR, NOT). For the population (workers), terms included controlled terms like "Workers" [MeSH], "Occupational Groups" [MeSH], "Employment" [MeSH], and free-text terms such as worker*, employee*, "working population", "labor force", occupation*, staff, and personnel. For exposure (irregular or rotating night shift work), controlled terms included "Shift Work Schedule" [MeSH], "Work Schedule Tolerance" [MeSH], "Circadian Rhythm" [MeSH], and free-text terms like "night shift*", "shift work", "rotating shift*", "irregular shift*", "quick return*", "short recovery", "frequent shift*", "shift change*", and "night work". Comparison terms (less frequent shifts, regular shifts, non-night work) included free-text terms such as "regular shift*", "fixed shift*", "non-rotating shift*", "day shift*", "permanent schedule*", "non-night shift*", and "standard work hours". For the outcome (migraine and non-subtyped headaches), controlled terms were "Migraine Disorders" [MeSH], "Headache" [MeSH], "Headache Disorders" [MeSH], and free-text terms included migraine*, headache*, cephalalgia, "head pain", and "cranial pain", with a note that subtypes like "tension-type" or "cluster" would be excluded unless part of broader headache data. Measurement and study design terms (odds ratios, relative risks, observational studies) included controlled terms like "Odds Ratio" [MeSH], "Relative Risk" [MeSH], "Observational Study" [MeSH], "Cross-Sectional Studies" [MeSH], "Case-Control Studies" [MeSH], "Cohort Studies" [MeSH], and free-text terms such as "odds ratio*", OR, prevalence, association, "cross-sectional", "case-control", cohort, observational, and epidemiology*.

An example search string for PubMed was: ("Workers"[MeSH Terms] OR "Occupational Groups"[MeSH Terms] OR "Employment"[MeSH Terms] OR worker* OR employee* OR "working population" OR occupation*) AND ("Shift Work Schedule"[MeSH Terms] OR "Work Schedule Tolerance"[MeSH Terms] OR "night shift*" OR "shift work" OR "rotating shift*" OR "irregular shift*" OR "quick return*" OR "night work") AND ("Migraine Disorders"[MeSH Terms] OR "Headache"[MeSH Terms] OR migraine* OR headache* OR cephalalgia) AND ("Odds Ratio"[MeSH Terms] OR "relative risk*" OR "hazard ratio*" OR "Observational Study"[MeSH Terms] OR "Cross-Sectional Studies"[MeSH Terms] OR "Case-Control Studies"[MeSH Terms] OR "Cohort Studies"[MeSH Terms] OR "odds ratio*" OR prevalence OR "cross-sectional" OR cohort OR observational). A similar structure was adapted for Web of Science: TS=((worker* OR employee* OR "working population" OR occupation*) AND ("night shift*" OR "shift work" OR "rotating shift*" OR "irregular shift*" OR "quick return*" OR "night work") AND (migraine* OR headache* OR cephalalgia) AND ("odds ratio*" OR "relative risk*" OR "hazard ratio*" OR prevalence OR "cross-sectional" OR "case-control" OR cohort OR observational)). Filters in PubMed included study type (Observational Study, Cross-Sectional Studies, Case-Control Studies, Cohort Studies where applicable), language (no restriction, translations sought if needed), and species (humans), while Web of Science filters included document type (Article, Conference Proceeding, Early Access), research area (Medicine, Public Environmental Occupational Health, Neurosciences, Epidemiology if applicable), and language (no restriction). There was no date restriction, with searches including all available records up to the current date (March 27, 2025), reflecting the lack of a cutoff in the inclusion criteria and the continuous knowledge update capability, which ensures comprehensive coverage of both historical and recent studies. Search strings, run dates, and result counts were recorded for transparency (Figure 1, PRISMA flow diagram). Reference lists of included studies and relevant reviews were manually searched for additional eligible studies. This strategy strikes a balance between sensitivity (capturing relevant studies) and specificity (focusing on the question’s scope). We prioritized ORs for consistency across study designs, although cohort studies often report relative risks or hazard ratios. Therefore, we included terms for these metrics in our search and calculated ORs where possible.

**Figure 1.**
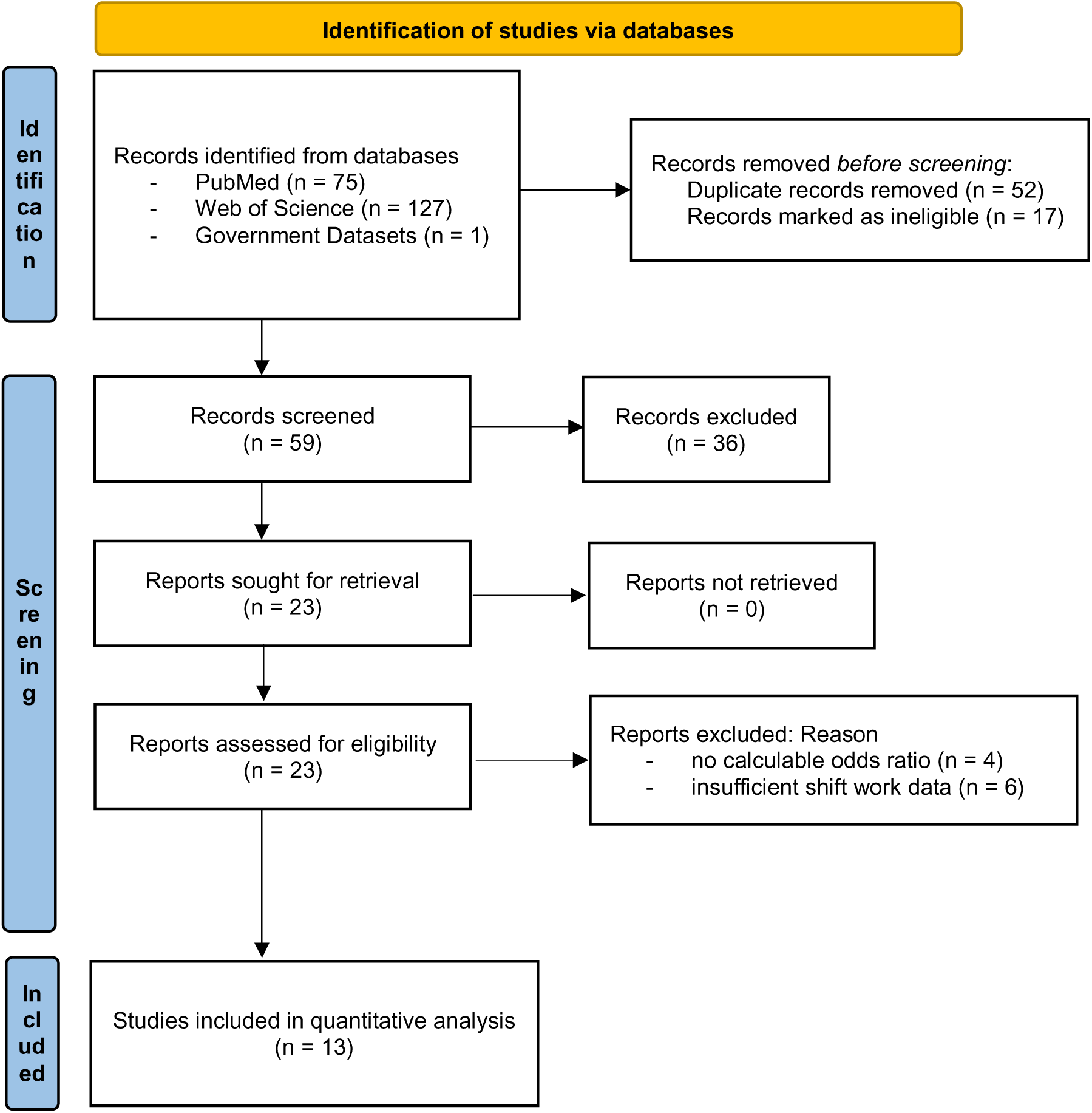
PRISMA Flow Diagram of Study Selection

### Data Extraction

The following data were extracted: first author, year of publication, country, sample size, male to female ratio, study design, type of comparison (rotating vs non-rotating night shiftwork, frequent vs infrequent irregular night shift work), and number of people with migraine or headache (where headache was not phenotyped) in the compared groups. Six authors participated in data extraction.

### Quality Assessment

The Newcastle Ottawa Scale^73^ (by YWW and MF) was used to assess the quality of each included article in the following three domains: selection of study groups, comparability of groups, and ascertainment of exposure or outcome.

### Statistical Analysis

A random-effects model was employed for the meta-analysis to account for between-study heterogeneity, which arises from differences in study settings, populations, and sample sizes. This model provides a more conservative estimate of the overall effect compared to a fixed-effects model. The inverse variance method with the DerSimonian-Laird estimator was used to weight studies based on their precision, accommodating the wide range of study sizes and heterogeneity effectively. In cases of sparse events, the robustness of this approach was considered, with the Mantel-Haenszel method as a potential alternative for sensitivity analyses to ensure stability in estimates. Odds ratios (ORs) were used as the effect measure instead of relative risk, as ORs provide consistent and comparable associations across observational studies, particularly cross-sectional designs. ORs are less sensitive to variations in baseline risks and are well-suited for studies where temporality or causality cannot be established. Data were synthesized using the online platform https://metaanalysisonline.com/^74^ to calculate effect sizes and assess heterogeneity. Meta-Essentials software^75^ was utilized to perform statistical analyses, generate forest plots, conduct meta-regression, and evaluate publication bias. All procedures adhered to standard meta-analysis guidelines to ensure robust and reliable results. The study protocol was registered on PROSPERO (CRD420250654865) on 24 February 2025. Inter-rater reliability checks and duplicate screening were conducted by YWW and EH to enhance methodological rigor.

## Results

Study Selection: Figure 1 presents the PRISMA flow diagram detailing the study selection process. A total of [n = 203] records were identified from PubMed ([n = 75]) and Web of Science ([n = 127]), as well as government dataset (NHIS). After removing [n = 69] duplicates, 59 records were screened by title and abstract, excluding [n = 36] as irrelevant. Of 23 full-text articles assessed for eligibility, 10 were excluded for reasons including insufficient shift work data (n = 6) or no odds ratios or calculable data (n = 4). Ultimately, 13 studies^46,76–87^ were included in quantitative synthesis.

Study Characteristics: The study analyzed a combined sample size of 38,798,271 participants, comprising 77% female participants. The participants originated from eight countries: China (3), Denmark (2), Saudi Arabia (2), Norway (1), UK (1), Singapore (1), Canada (1), USA (2). All included studies involved a cross-sectional design.

Quality Assessment (Table 1): The Newcastle-Ottawa Scale (NOS)^20^ scores for the listed studies range from 9 to 10, indicating high methodological quality across the board. The majority of studies (7 out of 12) scored a perfect score of 10, excelling in selection, comparability, and outcome criteria (Q1-Q7). The remaining five studies scored 9, with minor deductions primarily in Q2 or Q6, suggesting slight variations in representativeness or ascertainment of exposure and outcome.

**Table 1.** The Risk of Bias Assessment shows Newcastle-Ottawa Scale (NOS) scores ranging from 9 to 10, indicating a high methodological quality. Most studies (7 of 12) scored a perfect 10 in selection, comparability, and outcomes (Q1-Q7). The other five studies scored 9, with minor deductions mainly in Q2 or Q6, reflecting slight variations in representativeness or exposure/outcome ascertainment. The NHIS 2015 was not evaluated as it was a dataset.

|  | Study | Q1 | Q2 | Q3 | Q4 | Q5a | Q5b | Q6 | Q7 | Total |
| --- | --- | --- | --- | --- | --- | --- | --- | --- | --- | --- |
| 1 | Affatato 2021 | 1 | 1 | 1 | 2 | 1 | 1 | 2 | 1 | 10 |
| 2 | Al Maqwashi 2024 | 1 | 1 | 1 | 2 | 1 | 1 | 2 | 1 | 10 |
| 3 | Bjorvatn 2018 | 1 | 1 | 1 | 2 | 1 | 1 | 2 | 1 | 10 |
| 4 | Chan 1993 | 1 | 1 | 1 | 2 | 1 | 1 | 2 | 1 | 10 |
| 5 | Jacobsen 2017 | 1 | 1 | 1 | 2 | 1 | 1 | 2 | 1 | 10 |
| 6 | Jensen 2018 | 1 | 0 | 1 | 2 | 1 | 1 | 2 | 1 | 9 |
| 7 | Lees 1980 | 1 | 0 | 1 | 2 | 1 | 1 | 2 | 1 | 9 |
| 8 | Tasto 1978 | 1 | 0 | 1 | 2 | 1 | 1 | 2 | 1 | 9 |
| 9 | Wang 2015 | 1 | 1 | 1 | 2 | 1 | 1 | 2 | 1 | 10 |
| 10 | Xie 2020 | 1 | 0 | 1 | 2 | 1 | 1 | 2 | 1 | 9 |
| 11 | Alturaiki 2023 | 1 | 1 | 1 | 2 | 1 | 1 | 1 | 1 | 9 |
| 12 | Liu 2019 | 1 | 1 | 1 | 2 | 1 | 1 | 2 | 1 | 10 |

Quantitative Synthesis: Altogether, 13 studies were analyzed. Based on the analysis performed using a random effects model with the inverse variance method to compare the OR, a statistical difference was observed; the summarized OR was 1.61 with a 95% confidence interval of 1.27-2.04 (see Figure 2). The test for overall effect showed significance at p < 0.0001. The I^2^ value indicated a 73% inter-study heterogeneity.

**Figure 2.**
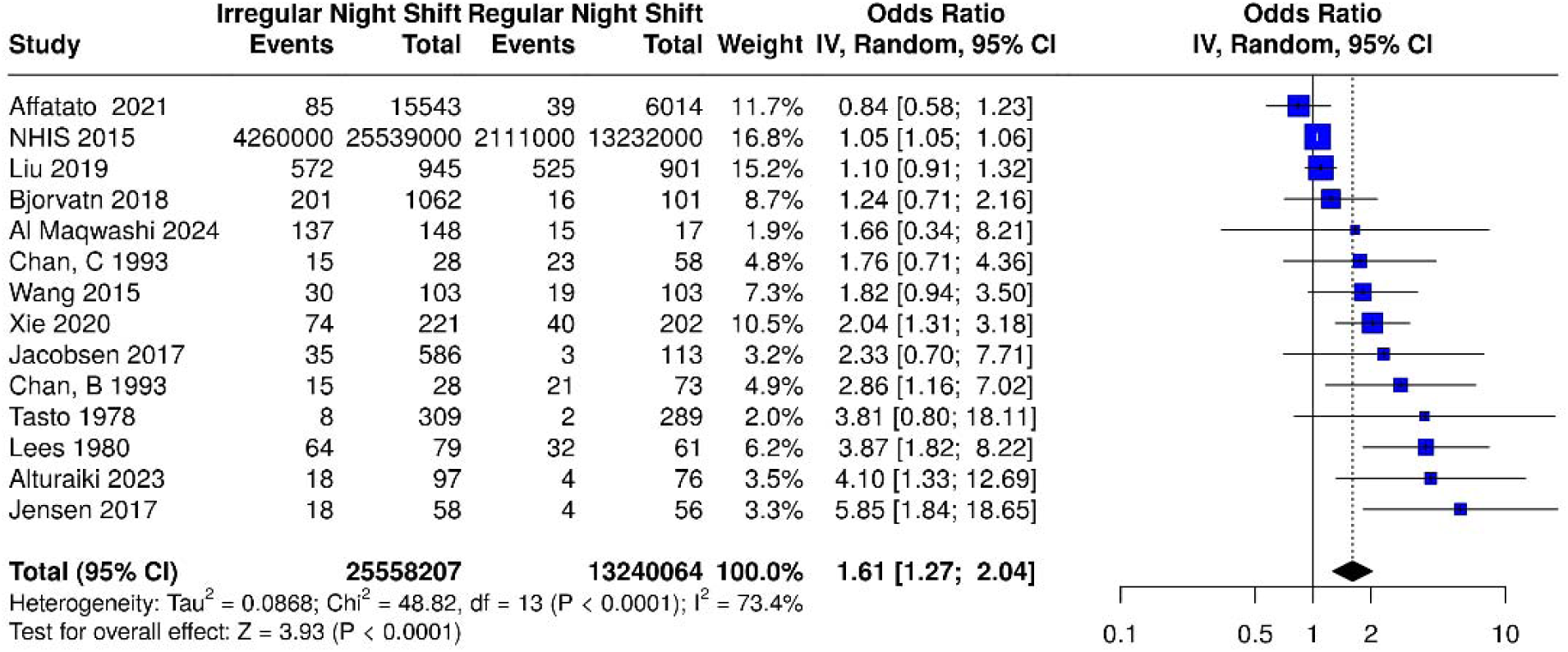
Based on the analysis performed using a random effects model with the inverse variance method to compare the odds ratio (OR), there was a statistically significant association between irregular night shift work and migraine prevalence; the summarized OR was 1.61 with a 95% confidence interval (CI) of 1.27 – 2.04 among the 13 studies (14 datasets) included. Abbreviations: IV = Inverse Variance; dF = degrees of freedom. Events represent the number of cases with migraine.

Among the 12 studies reviewed, sex-specific data were available in only one study^87^. An additional study, the only other in the literature providing sex-specific data^88^, was included, resulting in a total of 10,503 participants (45% female). The OR for females was 2.02 (95% CI: 1.71–2.39)^88^ in one study and 4.21 (95% CI: 2.09–8.46)^87^ in the other, suggesting that females had approximately 2–4 times higher odds of migraine associated with night shift work compared to males. A meta-regression across 12 studies, which provided male-to-female percentage data, reinforced this finding. The meta-regression analysis revealed a significant association between the proportion of females and increased migraine odds in irregular versus regular night shift workers, with a β (standardized beta) of 0.70 (p = 0.0003), accounting for 50% of the variance (Figure 3).

**Figure 3.**
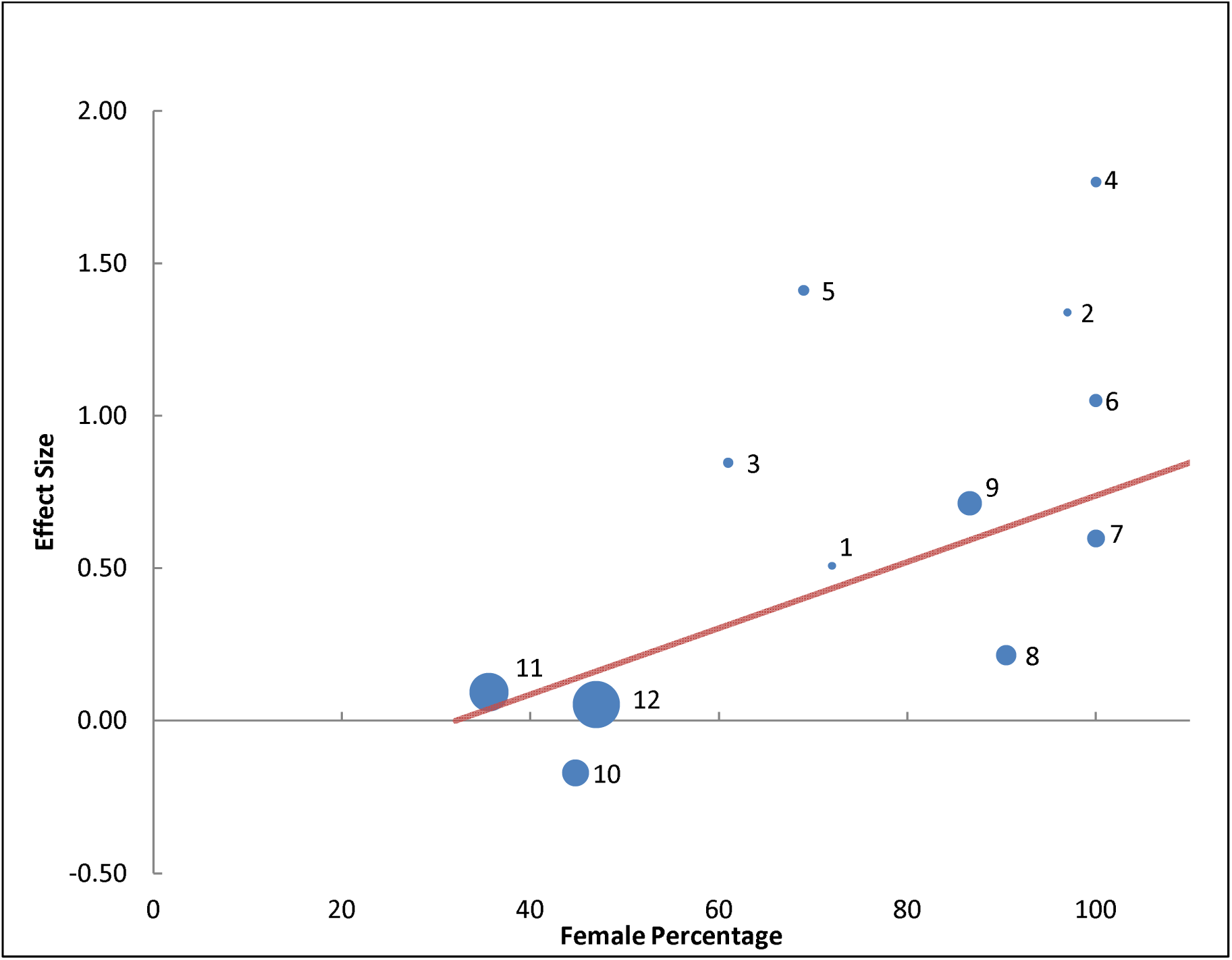
Meta-regression of 12 studies reporting male-to-female percentages, with bubble sizes indicating study weight and numbers corresponding to studies, showing a significant association between higher female proportion and increased migraine odds in irregular versus regular night shift workers (standardized β = 0.70, p = 0.0003), explaining 50% of the variance. Studies and their variance percentage contribution: 1 = Al Maqwashi 2024 (0.87%), 2 = Tasto 1978 (0.92%), 3 = Jacobsen 2017 (1.52%), 4 = Jensen 2017 (1.62%), 5 = Alturaiki 2023 (1.70%), 6 = Chan 1993 (2.61%), 7 = Wang 2015 (4.59%), 8 = Bjorvatn 2018 (6.05%), 9 = Xie 2020 (8.56%), 10 = Affatato 2021 (10.68%), 11 = Liu 2019 (21.94%), 12 = NHIS 2015 (32.80%).

A meta-analysis of four studies with available tension-type headache (TTH) data, conducted using a random-effects model with the inverse variance method, evaluated the association between irregular night shift work and TTH prevalence. The pooled OR was 0.79 (95% CI: 0.43– 1.45), indicating no statistically significant association (p > 0.05 for overall effect, I² = 80%; Figure 4). Subgroup analysis showed a significant effect for migraine-only (subgroup 1) and all headaches (including non-phenotyped, subgroup 2). No significant differences were observed between migraine-only and all headaches subgroups (p = 0.25), with migraine appearing as the primary driver of the overall effect.

**Figure 4.**
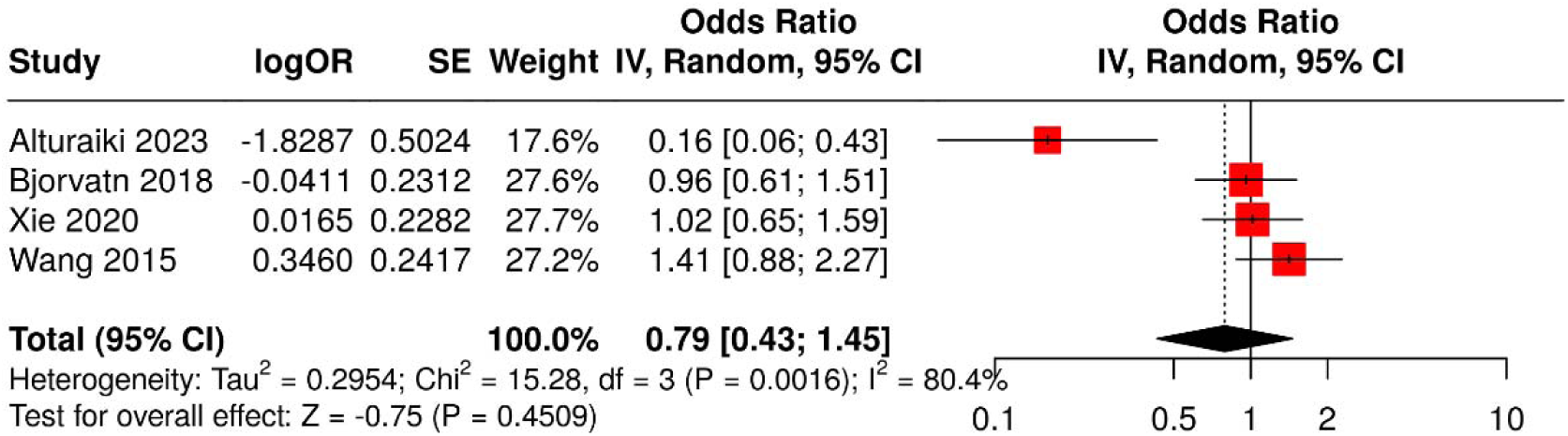
Meta-analysis of four studies using a random-effects model with inverse variance method to compare odds ratios (OR) of tension-type headache prevalence associated with irregular night shift work. The summarized OR was 0.79 (95% CI: 0.43–1.45), indicating no statistically significant association (test for overall effect, p > 0.05).

Meta-regression, using log-transformed (log10) sample size as a covariate to account for its wide range, showed no significant effect of sample size on the outcome (β = -0.31, p = 0.215).

## Discussion

Night shift work, a necessity in many professions, disrupts the delicate balance of the circadian system. This disruption leads to physiological stress, irregular sleep patterns, and heightened sensitivity to migraine triggers. Our meta-analytic evidence suggests a robust association between irregular or rotational shift work and a greater prevalence of migraine. Our findings confirm that night shift work, particularly its irregular scheduling, is associated with an increased migraine burden. This discussion elaborates on the implications of fixed versus rotating shift schedules, female susceptibility to night shift effects, introduces the concept of "Shift Work Migraine Disorder" (SWMD) as a potential subgroup within the migraine spectrum, necessitating tailored interventions, and proposes strategies to reduce migraine exacerbation.

## Fixed versus Irregular Shift Schedules

Our results align with prior evidence that fixed night shift schedules are preferable to irregular or rotating schedules for minimizing circadian disruption^70^. Irregular shifts, particularly those with rapid rotations (changing night shift schedules every 1–4 days^89^), quick returns (less than 11 hours of rest between shifts^90^), and consecutive night shifts (> 3 nights)^91,92^ desynchronize the SCN, the master circadian pacemaker, from peripheral clocks in organs such as the liver and gut^93^. This desynchronization manifests as irregular sleep-wake cycles, and gastrointestinal disturbances, and may amplify sensory processing^3,9,94,95^ to migraine triggers such as bright light, loud sound, and dietary irregularities, though direct evidence is needed. Fixed schedules, by contrast, allow for gradual circadian adaptation for workers transitioning from day to night shifts, involving a 12-hour schedule change, with the biological clock adjusting by approximately 1-2 hours per day^60,96^. Based on this adaptation rate, we recommend maintaining consistent shift schedules for at least 12-14 days to allow sufficient time for entrainment of the 12-hour shift, potentially reducing physiological stress and migraine risk, though further research is needed to confirm this effect.

Rotating shifts, when unavoidable, should follow a delay direction (morning → evening → night) rather than an advance direction (night → morning)^97–99^. This aligns with the natural tendency of the human circadian clock to delay rather than advance, as demonstrated in a meta-analysis, which found lower rates of sleep disruption and mood disturbances with delay-rotated schedules^97–99^. Additionally, minimizing consecutive night shifts—ideally to one per cycle— reduces cumulative sleep debt and cortisol dysregulation, both of which are implicated in migraine pathophysiology^100–104^.

Avoiding quick returns (shifts with <11 hours of rest) reduces fatigue and sleep disturbances, as short inter-shift intervals exacerbate circadian misalignment^90,105^. However, Katsfiraki et al. (2019) found no association between sleep duration and headaches in shift workers (OR = 1.00, 95% CI: 0.97–1.02), suggesting headache triggers may involve circadian factors rather than sleep loss^106^.

## Female Susceptibility to Night Shift Effects

The findings from our meta-regression as well as subgroup meta-analysis, encompassing a substantial sample of 10,503 individuals (45% female), reveal a significant sex-specific disparity in migraine risk associated with night shift work. Females working night shifts have more than twice the odds of experiencing migraine compared to their male counterparts. These findings are consistent with the female preponderance in migraine prevalence, which occupational factors like circadian disruption may intensify^50^. Although females are less likely to engage in night shift work than males (15.2% vs. 17.6% in non-daytime schedules)^46^, they face a higher burden of shift work-related migraine. These findings emphasize the importance of investigating the mechanisms underlying this sex difference, particularly in relation to the neurological consequences of shift work.

One plausible explanation for this disparity could be the inherent differences in circadian physiology between sexes, as females are known to have a shorter free-running circadian cycle compared to males^58,107,108^. This shorter intrinsic period—typically around 24.2 hours in females versus 24.5 hours in males^109^—may render females more vulnerable to the desynchronization caused by night shift work, potentially heightening migraine susceptibility. Hormonal fluctuations, more prominent in females due to menstrual cycles, pregnancy, or menopause, could compound this effect by interacting with circadian misalignment^110^. Additionally, females might experience greater sensitivity to sleep deprivation or stress—common among night shift workers—owing to neurobiological differences, such as altered circadian and stress response pathways^111,112^, potentially compounded by external factors like caregiving responsibilities^113,114^. The timing of stress exposure within the circadian cycle differentially impacts males and females^111,112^. Female mice display exaggerated stress response during their active phase, involving disruptions in clock gene expression^111^, which provides a potential mechanistic link to explain why female night shift workers might experience higher migraine prevalence. Women working night shifts are active during their natural resting period, potentially leading to a similar desynchronization of circadian rhythms and increased vulnerability to stressors, which could exacerbate or induce migraine attacks. Nonetheless, these findings underscore the need for further research into potential confounding factors such as lifestyle factors (e.g., mealtimes, exercise, light exposure, diet, caffeine intake) and genetic predispositions, as well as longitudinal studies to confirm causality. Exploring interventions, such as tailored shift schedules or circadian-aligned therapies, could offer promising avenues to reduce migraine risk among female night shift workers.

## Shift Work Migraine Disorder: A Proposed Subgroup

Our study highlights the association between irregular night shift work and increased migraine, suggesting that shift work-related migraine may warrant consideration as a distinct clinical phenomenon within the migraine spectrum, tentatively termed “Shift Work Migraine Disorder” (SWMD), and warrants further research. We hypothesize that chronic circadian misalignment in night shift workers amplifies trigeminovascular activation, a core mechanism of migraine^115^, through heightened neuroinflammation and oxidative stress^116–118^. Evidence from our meta-analysis—showing a significantly higher migraine prevalence in night shift workers vs day workers and irregular night shift workers versus fixed night workers—supports this notion. SWMD may be characterized by unique features, such as increased susceptibility to circadian misalignment triggers (e.g., light, sleep deprivation) and a refractory response to standard prophylactic treatments like topiramate or beta-blockers, necessitating specialized management.

Our meta-analysis found no significant association between irregular night shift work and tension-type headache (TTH) prevalence. SWMD’s specificity to migraine, rather than TTH, suggests that circadian disruption from irregular night shifts uniquely exacerbates migraine pathophysiology. This may involve heightened trigeminal sensitivity or disrupted sleep-wake cycles, which are more relevant to migraine triggers (e.g., bright lights, loud noises, dietary irregularities) than TTH. The lack of TTH association strengthens SWMD’s distinctiveness, as circadian misalignment appears to preferentially impact migraine-prone individuals.

Future research should validate SWMD through longitudinal studies to confirm its pathophysiology. If substantiated, SWMD could be incorporated into the International Classification of Headache Disorders, with proposed diagnostic criteria including:

A. Migraine Diagnosis: Meets ICHD-3^1^ criteria for migraine with or without aura.
B. Shift Work Exposure: History of irregular night shift work (e.g., rotating shifts^89^, quick returns^90^, or >3 consecutive night shifts^91,92^) for ≥3 months.
C. Temporal Association: Migraine onset or exacerbation temporally linked to shift work, with attacks occurring during or within 24 hours of shift changes.
D. Circadian Disruption Evidence: Documented sleep-wake irregularities (e.g., via actigraphy or sleep logs) or symptoms of circadian misalignment (e.g., excessive daytime sleepiness).
E. Exclusion of Other Causes: Symptoms not better explained by another headache disorder or medical condition.

Rationale: These criteria ensure the disorder is distinct from other migraine subtypes (e.g., chronic migraine defined by frequency or menstrual migraine linked to hormonal triggers) by tying it specifically with the environmental trigger of irregular night shift work and its underlying circadian pathophysiology. Analogous to Shift Work Sleep Disorder (SWSD)^119^, a recognized circadian rhythm sleep disorder, the proposed “Shift Work Migraine Disorder” may represent a distinct migraine subtype triggered by irregular night shift work’s circadian disruption, as evidenced by our study’s finding of increased migraine burden.

While we agree that sleep disruption is a recognized migraine trigger,^104,120^ we propose that SWMD is a distinct entity due to its specific association with the circadian misalignment inherent in shift work schedules. Unlike general sleep-related migraines, SWMD is characterized by a consistent temporal relationship between migraine attacks and shift transitions, supported by our meta-analysis. In contrast to Jet Lag Disorder, which is temporary and resolves with time^121^, SWSD’s longer-term nature and need for significant adaptation align more closely with SWMD’s chronicity in shift workers (Table 2). The unique occupational context and potential neuroendocrine mechanisms (e.g., melatonin suppression^72,122–124^) further differentiate SWMD. Formalizing SWMD as a distinct entity has critical implications for targeted interventions, night shift protocols, and workplace health policies, justifying its recognition as a novel headache disorder.

**Table 2.**
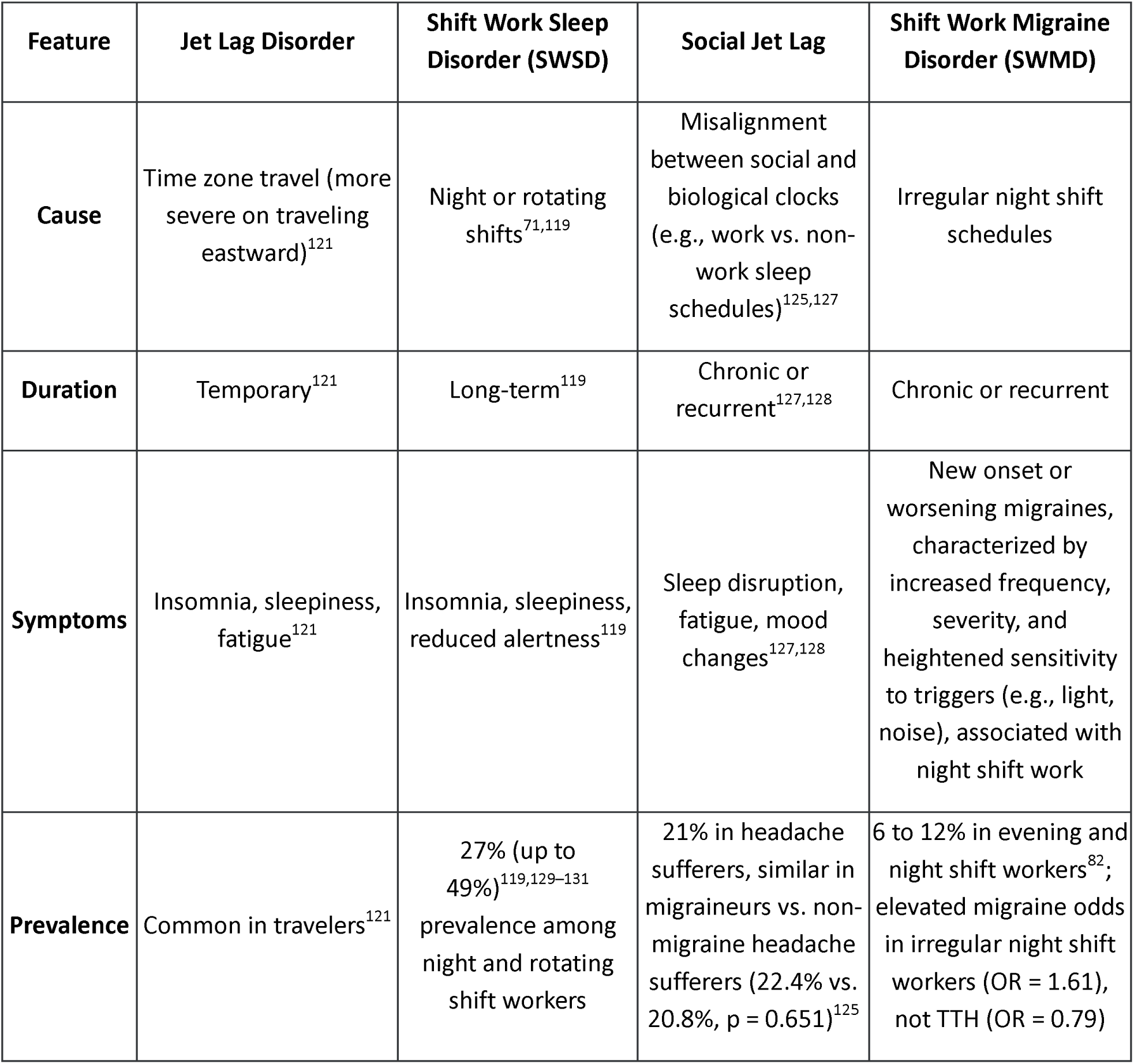
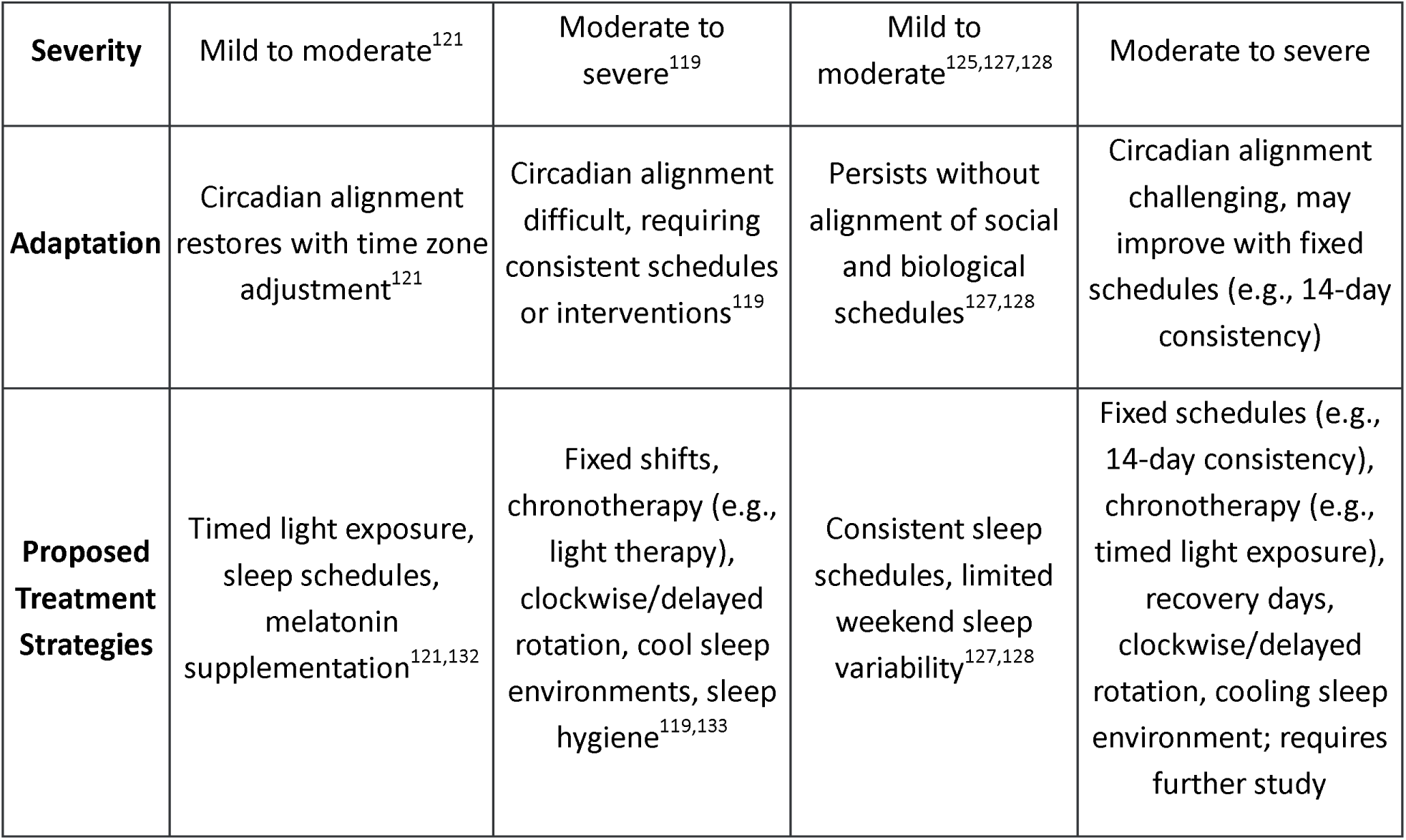
Comparison of Shift Work Related Conditions: Jet Lag Disorder, Shift Work Sleep Disorder (SWSD), Social Jet Lag, and proposed Shift Work Migraine Disorder (SWMD).

| Feature | Jet Lag Disorder | Shift Work Sleep Disorder (SWSD) | Social Jet Lag | Shift Work Migraine Disorder (SWMD) |
| --- | --- | --- | --- | --- |
| Cause | Time zone travel (more severe on traveling eastward)<sup>121</sup> | Night or rotating shifts<sup>71,119</sup> | Misalignment between social and biological clocks (e.g., work vs. non-work sleep schedules)<sup>125,127</sup> | Irregular night shift schedules |
| Duration | Temporary<sup>121</sup> | Long-term<sup>119</sup> | Chronic or recurrent<sup>127,128</sup> | Chronic or recurrent |
| Symptoms | Insomnia, sleepiness, fatigue<sup>121</sup> | Insomnia, sleepiness, reduced alertness<sup>119</sup> | Sleep disruption, fatigue, mood changes<sup>127,128</sup> | New onset or worsening migraines, characterized by increased frequency, severity, and heightened sensitivity to triggers (e.g., light, noise), associated with night shift work |
| Prevalence | Common in travelers<sup>121</sup> | 27% (up to 49%)<sup>119,129–131</sup> prevalence among night and rotating shift workers | 21% in headache sufferers, similar in migraineurs vs. non-migraine headache sufferers (22.4% vs. 20.8%, $p = 0.651$)<sup>125</sup> | 6 to 12% in evening and night shift workers<sup>82</sup>; elevated migraine odds in irregular night shift workers (OR = 1.61), not TTH (OR = 0.79) |

| **Severity** | Mild to moderate<sup>121</sup> | Moderate to severe<sup>119</sup> | Mild to moderate<sup>125,127,128</sup> | Moderate to severe |
| --- | --- | --- | --- | --- |
| **Adaptation** | Circadian alignment restores with time zone adjustment<sup>121</sup> | Circadian alignment difficult, requiring consistent schedules or interventions<sup>119</sup> | Persists without alignment of social and biological schedules<sup>127,128</sup> | Circadian alignment challenging, may improve with fixed schedules (e.g., 14-day consistency) |
| **Proposed Treatment Strategies** | Timed light exposure, sleep schedules, melatonin supplementation<sup>121,132</sup> | Fixed shifts, chronotherapy (e.g., light therapy), clockwise/delayed rotation, cool sleep environments, sleep hygiene<sup>119,133</sup> | Consistent sleep schedules, limited weekend sleep variability<sup>127,128</sup> | Fixed schedules (e.g., 14-day consistency), chronotherapy (e.g., timed light exposure), recovery days, clockwise/delayed rotation, cooling sleep environment; requires further study |

Table 2 compares Jet Lag Disorder, SWSD, Social Jet Lag, and the proposed SWMD, highlighting their distinct causes, durations, and symptoms. Unlike Jet Lag Disorder’s temporary effects^121^, SWMD’s chronicity and migraine-specific prevalence (elevated migraine odds with irregular night shift work, not TTH) align with SWSD’s long-term occupational impact, supporting SWMD as a novel headache disorder. Achieving circadian alignment can be challenging for SWMD and SWSD^119^, unlike Jet Lag Disorder, but fixed schedules (e.g., 14-day consistency) and chronotherapy may help, pending rigorous research. In a study of 2,762 participants, social jetlag was observed in 521 (18.9%) individuals and was more prevalent in headache sufferers (21.0%) than non-headache sufferers (17.0%, p=0.006)^125^. However, its prevalence did not significantly differ between migraineurs (22.4%) and non-migraine headache sufferers (20.8%, p=0.651)^125^. These findings suggest that social jetlag, a circadian misalignment, broadly affects headache disorders but does not uniquely distinguish migraines. In contrast, shift work migraine disorder, driven by occupational circadian disruption, may exhibit distinct clinical features, supporting its recognition as a novel entity separate from social jetlag and other circadian-related headache conditions in this large cohort^125^. These distinctions underscore the need for longitudinal studies to validate SWMD and its targeted interventions. The ongoing Phase 3 trial of solriamfetol (a dopamine and norepinephrine reuptake inhibitor approved for narcolepsy and obstructive sleep apnea) in SWSD (NCT06568367^126^) may offer insights for SWMD, as both conditions involve circadian disruption, but its efficacy for mitigating migraine burden requires rigorous investigation.

## Proposed Strategies: How to Work Night Shift and Mitigate Migraine Burden

Beyond shift scheduling^134^, our findings advocate for a multifaceted approach involving photic and non-photic circadian entrainment strategies to mitigate migraine burden in night shift workers. Potential management strategies for “Shift Work Migraine Disorder” could include chronotherapy (e.g., timed light exposure) and workplace policy changes, such as mandatory recovery days following shifts^134^, though their efficacy requires further investigation. Sleep health education is a cornerstone intervention, encompassing sleep hygiene (e.g., maintaining a cool, dark, quiet sleep environment), stimulus control (associating the bed with sleep only), and sleep restriction (limiting time in bed to actual sleep duration)^134^. A recent meta-analysis demonstrated that such interventions are efficacious in promoting sleep in night shift workers^134^. This highlights the need for further research into the relationship between circadian rhythms, night shift work, and migraine pathophysiology, with the goal of developing effective strategies to mitigate the risks associated with irregular work schedules.

### A. Photic Circadian Entrainment: Melanopic Lighting Interventions

Emerging evidence highlights the role of circadian or melanopic lighting in shift work environments^135–137^. Traditional fluorescent lighting, with low melanopic illuminance, fails to support circadian entrainment^138–140^. In contrast, dynamic lighting systems that mimic the natural day-night cycle—delivering high melanopic lux (e.g., 250-300 lux) during the shift and dim, warm light (e.g., <50 lux, 2700K) post-shift—have been shown to reduce fatigue and improve mood in night shift workers^140^. Such interventions could decrease migraine frequency by stabilizing circadian rhythms and reducing photic overstimulation. Implementing these systems in workplaces, although initially costly, may yield long-term benefits in terms of productivity and health outcomes.

Circadian resetting tools offer additional promise. The timing of light exposure, a potent zeitgeber, must be aligned with the minimum body temperature (Tmin), which typically occurs 2-3 hours before habitual wake time^141^. Mistimed light—particularly blue-rich light during the biological night—delays circadian adaptation and exacerbates photophobia^142^, a common migraine feature^1^. Blue-blocking glasses, worn during the latter half of night shifts, have been shown to reduce melatonin suppression and improve sleep quality^55^. Conversely, strategic exposure to bright, melanopic lighting (rich in 460-480 nm wavelengths) during the early shift can enhance alertness and facilitate circadian phase shifts^56^. In one study, light therapy glasses (461 nm, 22.34 μW/cm²) worn during night shifts significantly reduced driver sleepiness after the first night shift compared to placebo (p=0.012), supporting their use to enhance alertness and safety during commutes home for night shift workers^143^. This intervention, applied in a partial entrainment protocol, stabilizes sleepiness across night shifts^143^ and may mitigate migraine burden by aligning circadian rhythms with work schedules. In another study, blue-enriched white light (17,000 K, 89 lux) administered during night shifts improved subjective sleepiness compared to standard white light (4,000 K, 84 lux) when timed with the aMT6s peak, supporting its use to enhance alertness in night shift workers^137^ and potentially reduce migraine burden by mitigating circadian misalignment. Similarly, blue-enriched white light (>5,000 K) interventions significantly improved sleepiness in night-shift workers, including nurses, as demonstrated by a meta-analysis of 14 studies, supporting their use to enhance alertness^136^ and potentially reduce migraine burden by alleviating circadian desynchrony.

Based on circadian and light intervention studies involving night shift workers, we suggest reducing migraine burden by exposing workers to bright light (7,000–12,000 lux) before Tmin during night shifts^135^. This approach aims to cause a phase delay, aligning the circadian rhythm with a daytime sleep schedule, as evidenced by a study that shifted Tmin to midafternoon (14:53, p < 0.0001)^135^. Additionally, maintaining near-total darkness after Tmin—such as by using blue-blocking glasses during morning commutes—can help prevent phase advances that might interfere with adaptation^135^. Such circadian-informed light exposure shift protocols^144^ may reduce migraine frequency, severity, and trigger sensitivity by addressing circadian misalignment

### B. Non-photic Circadian Entrainment in Night Shift Workers

Non-photic entrainment methods for circadian adaptation in night shift workers include physical activity, meal timing, ambient temperature, social cues, and other behavioral strategies, which can be optimized with photic cues like light exposure. Physical activity, particularly light-to-moderate intensity exercise timed in the early evening (e.g., 19:00–22:00) or nocturnal hours (e.g., 00:30), can induce phase delays in circadian biomarkers like melatonin, improving alertness during night shifts and daytime sleep quality^145–147^. For instance, one hour of high-intensity nocturnal exercise can fully delay melatonin onset, aiding adaptation over a block of night shifts^145^. Meal timing, such as time-restricted eating within a 10-hour window aligned with the worker’s active period (e.g., during or shortly after night shifts), synchronizes metabolic and circadian rhythms, improves cognitive performance^148,149^, and reduces cardiometabolic risks like cardiovascular disease^150,151^. This approach avoids large late-night meals to minimize disruptions in glucose and insulin responses, as shown in a firefighter study^150^ and supported by evidence that large meals at 00:30 impair post-shift driving performance^151^.

Ambient temperature, another non-photic zeitgeber, affects circadian rhythms by adjusting body temperature, with cooler night environments potentially boosting alertness^152^. In one study, a "slightly cool" thermal sensation (around 23°C) enhanced work performance in night shift workers by increasing alertness and reducing thermal discomfort, which counters the sleep-inducing effects of circadian-driven sleepiness during night shifts^152^. Urinary melatonin levels decreased significantly during the second 23°C night shift, indicating improved circadian adaptation in night shift workers working in a cooler thermal environment^152^.

Social cues, such as scheduled interactions with colleagues or family during breaks or post-shift, provide temporal anchors to reinforce circadian entrainment^153–156^. These non-photic cues are most effective when combined with photic interventions^157^, like bright light during shifts and light avoidance post-shift (e.g., using blue-blocking glasses) to promote phase delays.

Pharmacological aids such as caffeine and melatonin also warrant consideration. Caffeine, when used judiciously (e.g., 100-200 mg at shift onset), boosts alertness. Caffeine, equivalent to two to four cups of coffee, significantly reduces physiological sleepiness and improves alertness during night shift hours, suggesting its potential use for critical night shift occupations. However, additional research is required to investigate how to optimize caffeine intake during night shifts, as it can impact daytime recovery sleep and sleep architecture^158^, considering caffeine’s well-known sleep-disrupting effects^159–161^.

Melatonin, administered at evidence-based doses (0.5-3 mg, 1-2 hours before desired sleep onset), accelerates circadian re-entrainment and reduces sleep onset latency, as confirmed in a recent systematic review^162,163^. A 2024 meta-analysis suggests that taking 4 mg of melatonin 3 hours before the desired bedtime may enhance its sleep-promoting effects compared to the commonly used regimen of 2 mg taken 30 minutes before bedtime, potentially due to better alignment with circadian phase-shifting needs^164^. This contrasts with the typical practice supported by earlier evidence^162,163^, where 1–3 mg taken 30–60 minutes before sleep effectively improves sleep onset and quality for shift workers, indicating that while the higher dose and earlier timing could optimize efficacy, the standard lower dose and shorter interval remain widely effective and practical. Timing is critical when using melatonin. In non-shift workers, evening melatonin induces a phase advance, shifting the circadian clock earlier, while morning melatonin causes a phase delay, shifting it later, aiding faster adaptation to new work shift schedules when timed appropriately^163,164^. Additionally, melatonin promotes sleep, indirectly influencing light-dark exposure by reducing retinal light input during sleep, further facilitating circadian clock resetting and improving sleep and alertness management^162,164^.

Limitations: The moderate heterogeneity observed in our meta-analysis is scientifically reasonable, given the diversity of the studies included. Our meta-analysis pools data from different working populations, countries, settings, and time periods. A consistent direction of effect across most studies (as seen in the forest plot, with the majority to the right) supports the observed trend.

## Recommendations and Implications

Drawing on evidence from circadian research, we suggest the following recommendations for night shift workers and employers, subject to further studies to verify their efficacy:

1. Shift Scheduling: Prioritize fixed night shifts or delay-rotated schedules, limit consecutive nights, and ensure ≥11-hour inter-shift intervals.
2. Sleep and Circadian Interventions: Implement sleep health education, use blue blockers and melanopic lighting, and time caffeine, melatonin, meals, and exercise to support entrainment.
3. Workplace Policies: Provide recovery time (e.g., 48 hours off after 3 consecutive nights) and consider a 14-day minimum shift consistency.
4. Research and Clinical Practice: Investigate SWMD’s mechanisms and trial chronotherapeutic protocols in affected populations.

If proven effective, these strategies could not only reduce migraine burden but also enhance overall well-being and productivity, potentially offering significant economic benefits by decreasing absenteeism and healthcare costs. Employers in healthcare, transportation, and manufacturing—sectors reliant on night shifts—stand to gain significantly from such interventions.

## Conclusion

Night shift work poses a formidable challenge to the general population, potentially triggering migraines or exacerbating pre-existing ones. Evidence-based scheduling and circadian management strategies, drawn from circadian research, could optimize adaptation, though these approaches require specific testing in migraine patients. Sleep or lighting interventions may help mitigate this burden, though their efficacy requires further study. A clockwise (morning, afternoon, night) shift rotation is less disruptive to the circadian rhythm than a counterclockwise (morning, night, afternoon) rotation, as humans adapt better to phase-delaying, and it provides longer rest periods between shifts^97,98^. Given the growing evidence linking circadian health to multiple chronic conditions, including migraine, recognizing ‘Shift Work Migraine Disorder’ (SWMD) as a potential clinical phenomenon is timely and warrants further investigation to improve the health of millions of night shift workers.

Conflicts of Interest: - YWW: Chief Medical Officer, Ex-Amplify Therapeutics; Consultant, Advanced Clinical and Research Center, Ethiopia; Consultant, Atheneum.

## Data Availability

Data supporting the findings of this study are available within the paper. Should any raw data files be needed in another format, they are available from the corresponding author upon reasonable request.

## Acknowledgements: None

Funding: This research was funded by the National Institute of Neurological Diseases and Stroke, National Institutes of Health, grant number K01NS124911 to YWW. The content is solely the responsibility of the authors and does not necessarily represent the official views of the National Institutes of Health.

## Ethical Approval and Informed Consent Statements

Not applicable.

## References

1. Headache Classification Committee of the International Headache Society (IHS) The International Classification of Headache Disorders, 3rd edition. Cephalalgia [online serial]. 2018;38:1–211. Accessed at: http://www.ncbi.nlm.nih.gov/pubmed/29368949.

2. Burstein R, Noseda R, Borsook D. Migraine: multiple processes, complex pathophysiology. J Neurosci [online serial]. 2015;35:6619–6629. Accessed at: http://www.ncbi.nlm.nih.gov/pubmed/25926442.

3. Ong JC, Taylor HL, Park M, et al. Can Circadian Dysregulation Exacerbate Migraines? Headache [online serial]. 2018;58:1040–1051. Accessed at: http://www.ncbi.nlm.nih.gov/pubmed/29727473.

4. Benkli B, Kim SY, Koike N, et al. Circadian Features of Cluster Headache and Migraine. Neurology. 2023;100.

5. Woldeamanuel YW, Palesh O, Cowan RP. Time it right! The Feasibility, Acceptability & Preliminary Efficacy of a Circadian-based Migraine Intervention. Ann Neurol. 2023;94:S152–S152.

6. Woldeamanuel YW, Xia C, Ding S, Fonteh A, Arakaki X. RNA-seq reveals transcriptomic differences in circadian-related genes of the choroid plexus in a preclinical chronic migraine model. bioRxiv. Epub 2025 Jun 6.

7. Gonzalez-Martinez A, Ray JC, Haghdoost F, et al. Time and headache: Insights into timing processes in primary headache disorders for diagnosis, underlying pathophysiology and treatment implications. Cephalalgia. 2024;44:3331024241297652.

8. Yang M-Y, Wu C-N, Lin Y-T, Tsai M-H, Hwang C-F, Yang C-H. Dissecting the Circadian Clock and Toll-like Receptor Gene Alterations in Meniere’s Disease and Vestibular Migraine. Otolaryngol Head Neck Surg. 2025;172:999–1005.

9. Poulsen AH, Younis S, Thuraiaiyah J, Ashina M. The chronobiology of migraine: a systematic review. J Headache Pain [online serial]. 2021;22:76. Accessed at: http://www.ncbi.nlm.nih.gov/pubmed/34281500.

10. Shirakawa Y, Ohno SN, Yamagata KA, et al. Circadian rhythm of PERIOD2::LUCIFERASE expression in the trigeminal ganglion of mice. Front Neurosci. 2023;17:1142785.

11. Schulte LH, May A. The migraine generator revisited: continuous scanning of the migraine cycle over 30 days and three spontaneous attacks. Brain. 2016;139:1987–1993.

12. Brennan KC, Bates EA, Shapiro RE, et al. Casein kinase iδ mutations in familial migraine and advanced sleep phase. Sci Transl Med. 2013;5:183ra56, 1–11.

13. Imai N. Molecular and Cellular Neurobiology of Circadian and Circannual Rhythms in Migraine: A Narrative Review. Int J Mol Sci. 2023;24.

14. Bumgarner JR, Walker WH, Nelson RJ. Circadian rhythms and pain. Neurosci Biobehav Rev. 2021;129:296–306.

15. Daguet I, Raverot V, Bouhassira D, Gronfier C. Circadian rhythmicity of pain sensitivity in humans. Brain. 2022;145:3225–3235.

16. Potter GDM, Skene DJ, Arendt J, Cade JE, Grant PJ, Hardie LJ. Circadian Rhythm and Sleep Disruption: Causes, Metabolic Consequences, and Countermeasures. Endocr Rev. 2016;37:584– 608.

17. Fishbein AB, Knutson KL, Zee PC. Circadian disruption and human health. J Clin Invest. 2021;131.

18. Kiehn J-T, Faltraco F, Palm D, Thome J, Oster H. Circadian Clocks in the Regulation of Neurotransmitter Systems. Pharmacopsychiatry. 2023;56:108–117.

19. Reinke H, Asher G. Crosstalk between metabolism and circadian clocks. Nat Rev Mol Cell Biol. 2019;20:227–241.

20. Innominato PF, Roche VP, Palesh OG, Ulusakarya A, Spiegel D, Lévi FA. The circadian timing system in clinical oncology. Ann Med. 2014;46:191–207.

21. Innominato PF, Komarzynski S, Palesh OG, et al. Circadian rest-activity rhythm as an objective biomarker of patient-reported outcomes in patients with advanced cancer. Cancer Med [online serial]. 2018;7:4396–4405. Accessed at: http://www.ncbi.nlm.nih.gov/pubmed/30088335.

22. IARC Monographs Vol 124 group. Carcinogenicity of night shift work. Lancet Oncol. 2019;20:1058–1059.

23. Amidi A, Wu LM. Circadian disruption and cancer- and treatment-related symptoms. Front Oncol. 2022;12:1009064.

24. Hadadi E, Taylor W, Li X-M, et al. Chronic circadian disruption modulates breast cancer stemness and immune microenvironment to drive metastasis in mice. Nat Commun. 2020;11:3193.

25. Zhang C, Tait C, Minacapelli CD, et al. The Role of Race, Sex, and Age in Circadian Disruption and Metabolic Disorders. Gastro Hep Advances. Elsevier; 2022;1:471–479.

26. Xu Y, Su S, McCall W V, Isales C, Snieder H, Wang X. Rest-activity circadian rhythm and impaired glucose tolerance in adults: an analysis of NHANES 2011–2014. BMJ Open Diabetes Res Care. 2022;10:e002632.

27. Peng X, Fan R, Xie L, et al. A Growing Link between Circadian Rhythms, Type 2 Diabetes Mellitus and Alzheimer’s Disease. Int J Mol Sci. 2022;23.

28. Warfield AE, Prather JF, Todd WD. Systems and Circuits Linking Chronic Pain and Circadian Rhythms. Front Neurosci. 2021;15.

29. Chellappa SL, Vujovic N, Williams JS, Scheer FAJL. Impact of Circadian Disruption on Cardiovascular Function and Disease. Trends Endocrinol Metab. 2019; 30:767–779.

30. Hoyt KR, Obrietan K. Circadian clocks, cognition, and Alzheimer’s disease: synaptic mechanisms, signaling effectors, and chronotherapeutics. Mol Neurodegener. 2022; 17:35.

31. Musiek ES, Bhimasani M, Zangrilli MA, Morris JC, Holtzman DM, Ju Y-ES. Circadian Rest-Activity Pattern Changes in Aging and Preclinical Alzheimer Disease. JAMA Neurol. 2018;75:582.

32. Ahmad F, Sachdeva P, Sarkar J, Izhaar R. Circadian dysfunction and Alzheimer’s disease - An updated review. Aging Med (Milton). 2023; 6:71–81.

33. Plain language about shiftwork. Rosa, Roger Rudolph; Colligan, Michael: National Institute for Occupational Safety and Health. Department of Health and Human Services. Division of Biomedical and Behavioral Science; Education and Information Division; Published Datelll: July 1997 Pages in Documentlll: print; vi, 38 p.lll: ill.lll; 28 cm. Serieslll: NIOSH Numbered Publications URLlll: https://stacks.cdc.gov/view/cdc/5177.

34. International Labour Organization. “Convention C171 - Night Work Convention, 1990 (No. 171).” NORMLEX, 26 June 1990, https://normlex.ilo.org/dyn/nrmlx/en/f?p=NORMLEXPUB:12100:0::NO:12100:P12100_INSTRUMENT_ID:312316:NO. Accessed 16 July 2025.

35. IARC Working Group on the Evaluation of Carcinogenic Risks to Humans. Painting, Firefighting, and Shiftwork. Lyon (FR): International Agency for Research on Cancer; 2010. (IARC Monographs on the Evaluation of Carcinogenic Risks to Humans, No. 98.) 1, Definition and Occurrence of Exposure. Available from: https://www.ncbi.nlm.nih.gov/books/NBK326824/.

36. European Union. “Directive 2003/88/EC of the European Parliament and of the Council of 4 November 2003 Concerning Certain Aspects of the Organisation of Working Time.” EUR-Lex, 4 Nov. 2003, https://eur-lex.europa.eu/legal-content/EN/TXT/PDF/?uri=CELEX:32003L0088. Accessed 16 July 2025.

37. Eurostat. “Employed Persons Working at Night by Sex, Age and Professional Status.” European Commission, 12 June 2025, https://ec.europa.eu/eurostat/databrowser/view/lfsa_ewpnig/default/line?lang=en&category=qoe.qoe_woli.qoe_wta. Accessed 16 July 2025.

38. Chile: Echeverría, M. Labour organization and time in Chile. ILO Conditions of Work and Employment Programme unpublished report, 2002.

39. PNAD. (2016). Síntese de indicadores. Coordenação de Trabalho e Rendimento. Rio de Janeiro, Brazil: Pesquisa Nacional por Amostra de Domicílios, Instituto Brasileiro de Geografia e Estatística (IBGE). [Portuguese].

40. Australian Bureau of Statistics (2012). Working time arrangements. Australia. November 2012. Report No. 6342.0. Canberra: Australia. Available from: https://www.abs.gov.au.

41. Zeng X, Liang LU, Idris SU. (2005). Working time in transition: the dual task of standardization and flexibilization in China, Conditions of Work and Employment Programme Series No. 11. Geneva, Switzerland: International Labour Office.

42. Lee S, McCann D, Messenger JC. (2007). Working time around the world. Trends in working hours, laws and policies in a global comparative perspective. Oxon: Routledge; and Geneva, Switzerland: International Labour Organization.

43. Ndiaye A (2006). Étude sur le temps de travail et l’organisation du travail au Sénégal. Conditions of Work and Employment Programme Series No. 13. Geneva, Switzerland: International Labour Office. [French].

44. Kubo T. Estimate of the Number of Night Shift Workers in Japan. J UOEH. 2014;36:273–276.

45. Williams C. Work-life balance of shift workers. Statistics Canada. https://www150.statcan.gc.ca/n1/en/pub/75-001-x/2008108/pdf/10677-eng.pdf?st=nnrihcKZ (Retrieved July 29, 2025). Epub 2008.

46. National Institute for Occupational Safety and Health. “Unadjusted Prevalence of Work Organization Characteristics (NHIS 2015) Among Workers (NHIS-OHS).” Centers for Disease Control and Prevention, 29 Oct. 2019, https://wwwn.cdc.gov/NIOSH-WHC/chart/ohs-workorg?T=OU&OU=*&V=R&chk_codes=False. Accessed 15 July 2025.

47. Stewart WF, Roy J, Lipton RB. Migraine prevalence, socioeconomic status, and social causation. Neurology. 2013; 81:948–955.

48. Lipton RB, Bigal ME, Diamond M, Freitag F, Reed ML, Stewart WF. Migraine prevalence, disease burden, and the need for preventive therapy. Neurology. 2007; 68:343–349.

49. Victor T, Hu X, Campbell J, Buse D, Lipton R. Migraine prevalence by age and sex in the United States: A life-span study. Cephalalgia. 2010; 30:1065–1072.

50. Woldeamanuel YW, Cowan RP. Migraine affects 1 in 10 people worldwide featuring recent rise: A systematic review and meta-analysis of community-based studies involving 6 million participants. J Neurol Sci [online serial]. Elsevier; 2017;372:307–315. Accessed at: https://linkinghub.elsevier.com/retrieve/pii/S0022510X16307742. Accessed December 7, 2016.

51. Bazargan M, Comini J, Kibe LW, Assari S, Cobb S. Association between Migraine and Quality of Life, Mental Health, Sleeping Disorders, and Health Care Utilization Among Older African American Adults. J Racial Ethn Health Disparities. 2024; 11:1530–1540.

52. Charleston L. Headache Disparities in African-Americans in the United States: A Narrative Review. J Natl Med Assoc [online serial]. 2021; 113:223–229. Accessed at: http://www.ncbi.nlm.nih.gov/pubmed/33160641.

53. Nicholson RA, Rooney M, Vo K, O’Laughlin E, Gordon M. Migraine care among different ethnicities: do disparities exist? Headache. 2006; 46:754–765.

54. Heckman BD, Holroyd KA, O’Donnell FJ, et al. Race differences in adherence to headache treatment appointments in persons with headache disorders. J Natl Med Assoc. 2008;100:247– 255.

55. Heckman BD, Merrill JC, Anderson T. Race, psychiatric comorbidity, and headache characteristics in patients in headache subspecialty treatment clinics. Ethn Health. 2013;18:34–52.

56. Getz M, Charleston L, Armand CE, Willis AW, Seng E. Perceived discrimination and migraine-specific quality of life: A cross-sectional survey study in a Black/African American sample. Headache. Epub 2025 Jun 2.

57. U.S. Bureau of Labor Statistics. Job Flexibilities and Work Schedules -- 2017-2018 Data from the American Time Use Survey. Washington, DC: U.S. Department of Labor, 2019. Accessed July 22, 2025. https://www.bls.gov/news.release/archives/flex2_09242019.pdf.

58. Eastman CI, Tomaka VA, Crowley SJ. Sex and ancestry determine the free-running circadian period. J Sleep Res [online serial]. 2017;26:547–550. Accessed at: http://www.ncbi.nlm.nih.gov/pubmed/28332253.

59. Eastman CI, Molina TA, Dziepak ME, Smith MR. Blacks (African Americans) have shorter free-running circadian periods than whites (Caucasian Americans). Chronobiol Int. 2012;29:1072– 1077.

60. Eastman CI, Suh C, Tomaka VA, Crowley SJ. Circadian rhythm phase shifts and endogenous free-running circadian period differ between African-Americans and European-Americans. Sci Rep. 2015;5:8381.

61. Heckman BD, Britton AJ. Headache in African Americans: An Overlooked Disparity. J Natl Med Assoc. 2015;107:39–45.

62. Heckman BD, Ellis G. Preventive medication adherence in African American and Caucasian headache patients. Headache. 2011;51:520–532.

63. Åkerstedt T, Narusyte J, Svedberg P. Night work, mortality, and the link to occupational group and sex. Scand J Work Environ Health. 2020;46:508–515.

64. Boivin DB, Boudreau P. Impacts of shift work on sleep and circadian rhythms. Pathol Biol (Paris). 2014;62:292–301.

65. Härmä M, Karhula K, Puttonen S, et al. Shift work with and without night work as a risk factor for fatigue and changes in sleep length: A cohort study with linkage to records on daily working hours. J Sleep Res. 2019;28:e12658.

66. Czeisler CA, Moore-Ede MC, Coleman RH. Rotating shift work schedules that disrupt sleep are improved by applying circadian principles. Science. 1982;217:460–463.

67. Schneider D, Harknett K. Consequences of Routine Work-Schedule Instability for Worker Health and Well-Being. Am Sociol Rev. 2019;84:82–114.

68. Vangelova K. The effect of shift rotation on variations of cortisol, fatigue and sleep in sound engineers. Ind Health. 2008;46:490–493.

69. Lin P-C, Chen C-H, Pan S-M, et al. The association between rotating shift work and increased occupational stress in nurses. J Occup Health. 2015;57:307–315.

70. Khan WAA, Jackson ML, Kennedy GA, Conduit R. A field investigation of the relationship between rotating shifts, sleep, mental health and physical activity of Australian paramedics. Sci Rep. 2021;11:866.

71. Kalmbach DA, Pillai V, Cheng P, Arnedt JT, Drake CL. Shift work disorder, depression, and anxiety in the transition to rotating shifts: the role of sleep reactivity. Sleep Med. 2015;16:1532–1538.

72. Zeitzer JM, Dijk DJ, Kronauer R, Brown E, Czeisler C. Sensitivity of the human circadian pacemaker to nocturnal light: melatonin phase resetting and suppression. J Physiol. 2000;526 Pt 3:695–702.

73. Wells, G. A., Shea, B., O’Connell, D., Peterson, J., Welch, V., Losos, M., & Tugwell, P. (n.d.). The Newcastle-Ottawa Scale (NOS) for assessing the quality of nonrandomised studies in meta-analyses. Ottawa Hospital Research Institute. Retrieved July 24, 2025, from https://www.ohri.ca/programs/clinical_epidemiology/oxford.asp.

74. Fekete JT, Győrffy B. MetaAnalysisOnline.com: Web-Based Tool for the Rapid Meta-Analysis of Clinical and Epidemiological Studies. J Med Internet Res. 2025;27:e64016.

75. Suurmond R, van Rhee H, Hak T. Introduction, comparison, and validation of Meta-Essentials: A free and simple tool for meta-analysis. Res Synth Methods. 2017;8:537–553.

76. Xie W, Li R, He M, et al. Prevalence and risk factors associated with headache amongst medical staff in South China. J Headache Pain. 2020;21:5.

77. Wang Y, Xie J, Yang F, et al. The prevalence of primary headache disorders and their associated factors among nursing staff in North China. J Headache Pain. 2015;16:4.

78. Tasto DL, Colligan MJ, Skjei EW, Polly SJ. Health Consequences of Shift Work. Report No. PB80176563. National Institute for Occupational Safety and Health. Epub 1978.

79. Liu H, Liu J, Chen M, et al. Sleep problems of healthcare workers in tertiary hospital and influencing factors identified through a multilevel analysis: a cross-sectional study in China. BMJ Open. 2019;9:e032239.

80. Lees RE, Romeril CS, Wetherall LD. A study of stress indicators in workers exposed to industrial noise. Can J Public Health. 1980;71:261–265.

81. Jensen HI, Larsen JW, Thomsen TD. The impact of shift work on intensive care nurses’ lives outside work: A cross-sectional study. J Clin Nurs. 2018;27:e703–e709.

82. Jakobsen GS, Timm AM, Hansen ÅM, Garde AH, Nabe-Nielsen K. The association between shift work and treatment-seeking migraine in Denmark. Ergonomics. 2017;60:1207–1217.

83. Chan OY, Gan SL, Yeo MH. Study on the health of female electronics workers on 12 hour shifts. Occup Med (Lond). 1993;43:143–148.

84. Bjorvatn B, Pallesen S, Moen BE, Waage S, Kristoffersen ES. Migraine, tension-type headache and medication-overuse headache in a large population of shift working nurses: a cross-sectional study in Norway. BMJ Open. 2018;8:e022403.

85. Alturaiki HM, Aldawood MA, Alghirash F, et al. Headache Characteristics and Risk Factors Among Healthcare Providers in Al-Ahsa, Saudi Arabia. Cureus. 2023;15:e45377.

86. Al Maqwashi LS, Sufyani AM, Bichara MM, et al. The Association Between Shift Work and Migraine Attacks Among Healthcare Workers in the Kingdom of Saudi Arabia. Cureus. 2024;16:e53315.

87. Affatato O, Miguet M, Schiöth HB, Mwinyi J. Major sex differences in migraine prevalence among occupational categories: a cross-sectional study using UK Biobank. J Headache Pain. 2021;22:145.

88. Molarius A, Tegelberg Å, Öhrvik J. Socio-Economic Factors, Lifestyle, and Headache Disorders — A Population-Based Study in Sweden. Headache: The Journal of Head and Face Pain. 2008;48:1426–1437.

89. Pilcher JJ, Lambert BJ, Huffcutt AI. Differential effects of permanent and rotating shifts on self-report sleep length: a meta-analytic review. Sleep. 2000;23:155–163.

90. Eldevik MF, Flo E, Moen BE, Pallesen S, Bjorvatn B. Insomnia, excessive sleepiness, excessive fatigue, anxiety, depression and shift work disorder in nurses having less than 11 hours in-between shifts. PLoS One. 2013;8:e70882.

91. James L, Elkins-Brown N, Wilson M, et al. The effects of three consecutive 12-hour shifts on cognition, sleepiness, and domains of nursing performance in day and night shift nurses: A quasi-experimental study. Int J Nurs Stud. 2021;123:104041.

92. Sim J, Yun B-Y, Lee J, et al. The Association Between the Number of Consecutive Night Shifts and Insomnia Among Shift Workers: A Multi-Center Study. Front Public Health. 2021;9.

93. Boivin DB, Boudreau P, Kosmadopoulos A. Disturbance of the Circadian System in Shift Work and Its Health Impact. J Biol Rhythms. 2022;37:3–28.

94. Daguet I, Bouhassira D, Demarquay G, Gronfier C. Visual discomfort and pain in migraine: homeostatic or circadian origin? [Doctoral dissertation]. Université Claude Bernard Lyon 1: HAL Theses; 2019. https://theses.hal.science/tel-04977136v1/file/TH2019DAGUETINES.pdf.

95. Strother L, Ptacek L, Goadsby P, Holland P. Environmental and Genetic Circadian Disruption Increases Migraine-Associated Phenotypes in Mice (2700). Neurology [online serial]. Wolters Kluwer; 2020;94:2700. Accessed at: 10.1212/WNL.94.15_supplement.2700.

96. Monk TH, Buysse DJ, Billy BD, DeGrazia JM. Using nine 2-h delays to achieve a 6-h advance disrupts sleep, alertness, and circadian rhythm. Aviat Space Environ Med. 2004;75:1049–1057.

97. Lavie P, Tzischinsky O, Epstein R, Zomer J. Sleep-wake cycle in shift workers on a “clockwise” and “counter-clockwise” rotation system. Isr J Med Sci. 1992;28:636–644.

98. Shiffer D, Minonzio M, Dipaola F, et al. Effects of Clockwise and Counterclockwise Job Shift Work Rotation on Sleep and Work-Life Balance on Hospital Nurses. Int J Environ Res Public Health. 2018;15.

99. Neil-Sztramko SE, Pahwa M, Demers PA, Gotay CC. Health-related interventions among night shift workers: a critical review of the literature. Scand J Work Environ Health. 2014;40:543–556.

100. Vetter C, Fischer D, Matera JL, Roenneberg T. Aligning work and circadian time in shift workers improves sleep and reduces circadian disruption. Curr Biol. 2015;25:907–911.

101. Woldeamanuel YW, Sanjanwala BM, Cowan RP. Endogenous glucocorticoids may serve as biomarkers for migraine chronification. Ther Adv Chronic Dis [online serial]. 2020;11:204062232093979. Accessed at: https://journals.sagepub.com/doi/10.1177/2040622320939793.

102. Mackus M, Kraneveld A, Garssen J, Verster J. 1161 MIGRAINE AND SLEEP: A BI-DIRECTIONAL ASSOCIATION. Sleep [online serial]. 2017;40:A433–A433. Accessed at: https://academic.oup.com/sleep/article-lookup/doi/10.1093/sleepj/zsx050.1160.

103. Lin Y-K, Lin G-Y, Lee J-T, et al. Associations Between Sleep Quality and Migraine Frequency: A Cross-Sectional Case-Control Study. Medicine [online serial]. 2016;95:e3554. Accessed at: http://www.ncbi.nlm.nih.gov/pubmed/27124064.

104. Kelman L, Rains JC. Headache and sleep: examination of sleep patterns and complaints in a large clinical sample of migraineurs. Headache [online serial]. 2005;45:904–910. Accessed at: http://www.ncbi.nlm.nih.gov/pubmed/15985108.

105. Akerstedt T, Wright KP. Sleep Loss and Fatigue in Shift Work and Shift Work Disorder. Sleep Med Clin. 2009;4:257–271.

106. Katsifaraki M, Nilsen KB, Christensen JO, et al. Sleep duration mediates abdominal and lower-extremity pain after night work in nurses. Int Arch Occup Environ Health. 2019;92:415–422.

107. Crowley SJ, Eastman CI. Free-running circadian period in adolescents and adults. J Sleep Res. 2018;27.

108. Crowley SJ, Acebo C, Carskadon MA. Sleep, circadian rhythms, and delayed phase in adolescence. Sleep Med. 2007;8:602–612.

109. Duffy JF, Cain SW, Chang A-M, et al. Sex difference in the near-24-hour intrinsic period of the human circadian timing system. Proceedings of the National Academy of Sciences. 2011;108:15602–15608.

110. Chau YM, West S, Mapedzahama V. Night Work and the Reproductive Health of Women: An Integrated Literature Review. J Midwifery Womens Health. 2014;59:113–126.

111. Mingardi J, Giovenzana M, Nicosia N, Misztak P, Ieraci A, Musazzi L. Sex and Circadian Rhythm Dependent Behavioral Effects of Chronic Stress in Mice and Modulation of Clock Genes in the Prefrontal Cortex. Int J Mol Sci. 2025;26:6410.

112. Verma P, Hellemans KGC, Choi FY, Yu W, Weinberg J. Circadian phase and sex effects on depressive/anxiety-like behaviors and HPA axis responses to acute stress. Physiol Behav. 2010;99:276–285.

113. DePasquale N, Sliwinski MJ, Zarit SH, Buxton OM, Almeida DM. Unpaid Caregiving Roles and Sleep Among Women Working in Nursing Homes: A Longitudinal Study. Gerontologist. 2019;59:474–485.

114. Korompeli A, Chara T, Chrysoula L, Sourtzi P. Sleep Disturbance in Nursing Personnel Working Shifts. Nurs Forum (Auckl). 2013;48:45–53.

115. Ashina M, Hansen JM, Do TP, Melo-Carrillo A, Burstein R, Moskowitz MA. Migraine and the trigeminovascular system—40 years and counting. Lancet Neurol. 2019;18:795–804.

116. Chen Q-W, Meng R-T, Ko C-Y. Modulating oxidative stress and neurogenic inflammation: the role of topiramate in migraine treatment. Front Aging Neurosci. 2024;16.

117. Borkum JM. Migraine Triggers and Oxidative Stress: A Narrative Review and Synthesis. Headache: The Journal of Head and Face Pain. 2016;56:12–35.

118. Han C, Lim JY, Koike N, et al. Regulation of headache response and transcriptomic network by the trigeminal ganglion clock. Headache [online serial]. 2024;64:195–210. Accessed at: http://www.ncbi.nlm.nih.gov/pubmed/38288634.

119. Sachdeva A, Goldstein C. Shift Work Sleep Disorder. Circadian Rhythm Sleep-Wake Disorders. Cham: Springer International Publishing; 2020. p. 149–182.

120. Woldeamanuel YW, Cowan RP. The impact of regular lifestyle behavior in migraine: a prevalence case–referent study. J Neurol. 2016;263.

121. Avidan A, Colwell C. Jet lag syndrome: circadian organization, pathophysiology, and management strategies. Nat Sci Sleep. Epub 2010 Aug.:187.

122. Wei T, Li C, Heng Y, et al. Association between night-shift work and level of melatonin: systematic review and meta-analysis. Sleep Med. 2020;75:502–509.

123. Hunter CM, Figueiro MG. Measuring Light at Night and Melatonin Levels in Shift Workers: A Review of the Literature. Biol Res Nurs. 2017;19:365–374.

124. Lowden A, Åkerstedt T, Wibom R. Suppression of sleepiness and melatonin by bright light exposure during breaks in night work. J Sleep Res. 2004;13:37–43.

125. Chu M, Cho S, Joo E, Kim J. Social jetlag is prevalent among headache sufferers: a population-based study [abstract PF49]. . Headache. 2015;55:171.

126. Axsome Therapeutics, Inc. (2025). Studying solriamfetol modulation of TAAR-1, dopamine, and norepinephrine in shift work disorder (SUSTAIN) (NCT06568367). ClinicalTrials.gov. Retrieved July 29, 2025, from https://clinicaltrials.gov/study/NCT06568367.

127. Wittmann M, Dinich J, Merrow M, Roenneberg T. Social Jetlag: Misalignment of Biological and Social Time. Chronobiol Int. 2006;23:497–509.

128. Caliandro R, Streng AA, van Kerkhof LWM, van der Horst GTJ, Chaves I. Social Jetlag and Related Risks for Human Health: A Timely Review. Nutrients. 2021;13:4543.

129. Li Y, Lv X, Li R, et al. Predictors of Shift Work Sleep Disorder Among Nurses During the COVID-19 Pandemic: A Multicenter Cross-Sectional Study. Front Public Health. 2021;9.

130. Pallesen S, Bjorvatn B, Waage S, Harris A, Sagoe D. Prevalence of Shift Work Disorder: A Systematic Review and Meta-Analysis. Front Psychol. 2021;12.

131. Drake CL, Roehrs T, Richardson G, Walsh JK, Roth T. Shift Work Sleep Disorder: Prevalence and Consequences Beyond that of Symptomatic Day Workers. Sleep. 2004;27:1453–1462.

132. Eastman CI, Burgess HJ. How to Travel the World Without Jet Lag. Sleep Med Clin. 2009;4:241– 255.

133. Drake CL, Wright KP. Chapter 71 - Shift Work, Shift-Work Disorder, and Jet Lag. In: Kryger MH, Roth T, Dement WC, editors. Principles and Practice of Sleep Medicine (Fifth Edition) [online]. Philadelphia: W.B. Saunders; 2011. p. 784–798. Accessed at: https://www.sciencedirect.com/science/article/pii/B9781416066453000712.

134. Jeon BM, Kim SH, Shin SH. Effectiveness of sleep interventions for rotating night shift workers: a systematic review and meta-analysis. Front Public Health. 2023;11.

135. Czeisler CA, Johnson MP, Duffy JF, Brown EN, Ronda JM, Kronauer RE. Exposure to Bright Light and Darkness to Treat Physiologic Maladaptation to Night Work. New England Journal of Medicine. 1990;322:1253–1259.

136. Wu C-J, Huang T-Y, Ou S-F, Shiea J-T, Lee B-O. Effects of Lighting Interventions to Improve Sleepiness in Night-Shift Workers: A Systematic Review and Meta-Analysis. Healthcare. 2022;10:1390.

137. Sletten TL, Ftouni S, Nicholas CL, et al. Randomised controlled trial of the efficacy of a blue-enriched light intervention to improve alertness and performance in night shift workers. Occup Environ Med. 2017;74:792–801.

138. Cain SW, McGlashan EM, Vidafar P, et al. Evening home lighting adversely impacts the circadian system and sleep. Sci Rep. 2020;10:19110.

139. National Toxicology Program. NTP Cancer Hazard Assessment Report on Night Shift Work and Light at Night. Research Triangle Park (NC): National Toxicology Program; 2021 Apr. 2, Light at Night and Night Shift Work: Circadian Disruption Studies. Available from: https://www.ncbi.nlm.nih.gov/books/NBK571600/.

140. Nie J, Zhou T, Chen Z, et al. The effects of dynamic daylight-like light on the rhythm, cognition, and mood of irregular shift workers in closed environment. Sci Rep. 2021;11:13059.

141. Mellette HC, Hutt BK, Askovitz SI, Horvath SM. Diurnal Variations in Body Temperatures. J Appl Physiol. 1951;3:665–675.

142. Chellappa SL. Individual differences in light sensitivity affect sleep and circadian rhythms. Sleep. 2021;44.

143. Aarts MPJ, Hartmeyer SL, Morsink K, Kort HSM, de Kort YAW. Can Special Light Glasses Reduce Sleepiness and Improve Sleep of Nightshift Workers? A Placebo-Controlled Explorative Field Study. Clocks Sleep. 2020;2:225–245.

144. Guyett A, Lovato N, Manners J, et al. A circadian-informed lighting intervention accelerates circadian adjustment to a night work schedule in a submarine lighting environment. Sleep. 2024;47.

145. Buxton OM, Lee CW, L’Hermite-Balériaux M, Turek FW, Van Cauter E. Exercise elicits phase shifts and acute alterations of melatonin that vary with circadian phase. American Journal of Physiology-Regulatory, Integrative and Comparative Physiology. 2003;284:R714–R724.

146. Youngstedt SD, Elliott JA, Kripke DF. Human circadian phase-response curves for exercise. J Physiol. 2019;597:2253–2268.

147. Youngstedt SD, Kline CE, Elliott JA, Zielinski MR, Devlin TM, Moore TA. Circadian Phase-Shifting Effects of Bright Light, Exercise, and Bright Light + Exercise. J Circadian Rhythms. 2016;14:2.

148. Gupta CC, Centofanti S, Dorrian J, et al. Altering meal timing to improve cognitive performance during simulated nightshifts. Chronobiol Int. 2019;36:1691–1713.

149. Gupta CC, Dorrian J, Grant CL, et al. It’s not just what you eat but when: The impact of eating a meal during simulated shift work on driving performance. Chronobiol Int. 2017;34:66–77.

150. Manoogian ENC, Zadourian A, Lo HC, et al. Feasibility of time-restricted eating and impacts on cardiometabolic health in 24-h shift workers: The Healthy Heroes randomized control trial. Cell Metab. 2022;34:1442–1456.e7.

151. Gupta CC, Centofanti S, Dorrian J, et al. The impact of a meal, snack, or not eating during the night shift on simulated driving performance post-shift. Scand J Work Environ Health. 2021;47:78–84.

152. Kim JH, Song Y. The effects of indoor ambient temperature at work on physiological adaptation in night shift nurses. J Nurs Manag. 2020;28:1098–1103.

153. Aschoff J, Fatranska M, Giedke H, Doerr P, Stamm D, Wisser H. Human Circadian Rhythms in Continuous Darkness: Entrainment by Social Cues. Science (1979). 1971;171:213–215.

154. Easton DF, Gupta CC, Vincent GE, Ferguson SA. Move the night way: how can physical activity facilitate adaptation to shift work? Commun Biol. 2024;7:259.

155. Mistlberger RE, Skene DJ. Social influences on mammalian circadian rhythms: animal and human studies. Biological Reviews. 2004;79:533–556.

156. Elmore SK, Betrus PA, Burr R. Light, social zeitgebers, and the sleep–wake cycle in the entrainment of human circadian rhythms. Res Nurs Health. 1994;17:471–478.

157. Mistlberger RE, Skene DJ. Nonphotic Entrainment in Humans? J Biol Rhythms. 2005;20:339–352.

158. Carrier J, Fernandez-Bolanos M, Robillard R, et al. Effects of Caffeine are more Marked on Daytime Recovery Sleep than on Nocturnal Sleep. Neuropsychopharmacology. 2007;32:964– 972.

159. McHill AW, Smith BJ, Wright KP. Effects of Caffeine on Skin and Core Temperatures, Alertness, and Recovery Sleep During Circadian Misalignment. J Biol Rhythms. 2014;29:131–143.

160. Walsh JK, Muehlbach MJ, Humm TM, Stokes Dickins Q, Sugerman JL, Schweitzer PK. Effect of caffeine on physiological sleep tendency and ability to sustain wakefulness at night. Psychopharmacology (Berl). 1990;101:271–273.

161. Institute of Medicine (US) Committee on Military Nutrition Research; Marriott BM, editor. Food Components to Enhance Performance: An Evaluation of Potential Performance-Enhancing Food Components for Operational Rations. Washington (DC): National Academies Press (US); 1994. 20, Effects of Caffeine on Cognitive Performance, Mood, and Alertness in Sleep-Deprived Humans. Available from: https://www.ncbi.nlm.nih.gov/books/NBK209050/#.

162. Hannemann J, Laing A, Middleton B, et al. Effect of oral melatonin treatment on insulin resistance and diurnal blood pressure variability in night shift workers. A double-blind, randomized, placebo-controlled study. Pharmacol Res. 2024;199:107011.

163. Carriedo-Diez B, Tosoratto-Venturi JL, Cantón-Manzano C, Wanden-Berghe C, Sanz-Valero J. The Effects of the Exogenous Melatonin on Shift Work Sleep Disorder in Health Personnel: A Systematic Review. Int J Environ Res Public Health. 2022;19:10199.

164. Cruz-Sanabria F, Bruno S, Crippa A, et al. Optimizing the Time and Dose of Melatonin as a Sleep-Promoting Drug: A Systematic Review of Randomized Controlled Trials and Dose−Response Meta-Analysis. J Pineal Res. 2024;76.

